# SARS-CoV-2 Aggregated Activity Level Across Ontario Canada, Measured with the US CDC Wastewater Viral Activity Level (WVAL) Metric

**DOI:** 10.1101/2025.02.07.25321887

**Authors:** Robert Delatolla, Christopher T. DeGroot, Robert Michael McKay, Mark R. Servos, Steven McAvoy, Sanjena Krishnakumar, Orchid Jo, Michael Fung, Sherif Hegazy, Tim Fletcher, Albert Simhon, Vince Pileggi

**Affiliations:** University of Ottawa, 75 Laurier Avenue East, Ottawa, K1N 6N5, Ontario, Canada; University of Western Ontario, 1151 Richmond Street, London, N6A 3K7, Ontario, Canada; University of Windsor, 401 Sunset Avenue, Windsor, N9B 3P4, Ontario, Canada; University of Waterloo, 200 University Avenue West, Waterloo, N2L 3G1, Ontario, Canada; Ontario Ministry of the Environment, Conservation and Parks, 4th Floor, 40 St. Clair Avenue West, Toronto, M7A 2J3, Ontario, Canada; Retired, formerly with the Ontario Ministry of the Environment, Conservation and Parks, 4th Floor, 40 St. Clair Avenue West, Toronto, M7A 2J3, Ontario, Canada

**Author notes:** Corresponding author (Robert Delatolla), (Vince Pileggi).

**Keywords:** *Keywords*: Wastewater viral activity level, Wastewater and environmental surveillance, Wastewater-based epidemiology, Aggregation, Correlation with clinical cases, Normalization

## Abstract

The wastewater viral activity level (WVAL) was developed by the United States Centers for Disease Control and Prevention (US CDC) as a standardized metric to aggregate SARS-CoV-2 wastewater data, enabling the assessment of infection levels and trends at state/territorial, regional and national scales. This approach also facilitates comparative analysis of SARS-CoV-2 prevalence across regions. In this study, we developed and evaluated graphical methods to integrate the WVAL metric into interpretable visualizations for public health decision-making. Preliminary analysis demonstrated that WVAL values correlated strongly with clinical case counts, supporting its role as a confirmatory epidemiological measure. The WVAL framework provided a linear quantification method, allowing for the comparison of regional variations in infection patterns. This study leveraged data from the Ontario Wastewater Surveillance Initiative (Ontario WSI), which included over 100 sampling sites across seven geographical regions. Weekly mean WVAL values were computed for each site and aggregated at regional and provincial levels. In total, 59 sites contributed to the provincial WVAL calculation. The computational aggregation method followed the US CDC’s WVAL approach and was generally comparable to the Public Health Ontario (PHO) aggregation method, with the notable improvement of incorporating a linear level scale. Overall, this study demonstrated that WVAL effectively quantified SARS-CoV-2 differences at a public health regional scale. The WVAL metric proved to be a robust epidemiological tool, complementing other surveillance measures to support public health decision-making.

## Introduction

During the COVID-19 pandemic, wastewater surveillance emerged as a critical tool to support public health in tracking trends in community infections [1–7]. Many jurisdictions around the world quickly implemented wastewater surveillance, including Ontario, Canada, where a network of more than 100 sites was monitored for several years [8, 9]. At many sites across Ontario, strong correlations were observed between wastewater SARS-CoV-2 signals and clinical case trends [3, 10]. The Wastewater Surveillance Initiative (WSI) was established in 2020 to coordinate wastewater surveillance efforts and consistently report findings to public health agencies. However, aggregating and interpreting wastewater data regionally and temporally remained a challenge.

The Ontario COVID-19 Science Advisory Table [1] developed an aggregation method based on a weighted mean of biomarker-normalized viral concentrations, later adopted by Public Health Ontario (PHO) [11] for regional reporting. At the national level, the Public Health Agency of Canada (PHAC) [12] reports wastewater viral loads using a viral activity index (WVI)—a pseudo- aggregation method that classifies viral activity (low to high) based on historical percentiles but lacks a standardized quantitative scale for cross-region comparisons.

To address these aggregation inconsistencies, the United States Centers for Disease Control and Prevention (US CDC) [13] developed the Wastewater Viral Activity Level (WVAL) metric, which introduces a standardized method for comparing wastewater SARS-CoV-2 levels across jurisdictions. Unlike most pseudo-aggregation methods, WVAL applies log-normalization and statistical scaling, enabling direct quantitative comparisons across spatial and temporal boundaries.

Globally, many countries have implemented wastewater surveillance but rely on trend-based pseudo-aggregation methods rather than a standardized metric like WVAL. The National Institute for Public Health (RIVM) [14] in the Netherlands aggregates viral mass loading adjusted for population but lacks a fecal indicator or standardized scale. The Federal Environment Agency of Germany [15] normalizes viral concentrations using wastewater inflow ratios, applying log transformation and LOESS smoothing, but without a comparative scale. The VATar COVID-19 Spanish System [16] normalizes viral RNA by population and flow, integrating wastewater data with epidemiological trends, but without a fixed aggregation metric. The Obépine Network in France [17] measures wastewater SARS-CoV-2 levels with population and flow normalization but focuses on trend analysis rather than a standardized viral activity scale. The National Institute of Health in Italy [18] applies flow-adjusted normalization and correlates wastewater data with epidemiological models but lacks a unified comparison scale. The Finnish Institute for Health and Welfare (THL) [19] classifies viral loads as low, moderate, or high based on historical trends, integrating genomic sequencing rather than a fixed numerical scale. The Austrian Agency for Health and Food Safety [20] categorizes viral activity into increasing, decreasing, or stable trends, using a method akin to the epidemiological R-value, but without a standardized WVAL-like scale. Sciensano, Belgium’s public health institute [21], applies a trend-based classification system (e.g., “high circulation”) rather than absolute viral activity levels. The Portuguese Environment Agency [22] normalizes viral RNA using flow and population adjustments but focuses on trend analysis rather than a standardized scale. The Statens Serum Institut (SSI) of Denmark [23] uses trend-based classifications and integrates genomic sequencing to track SARS-CoV-2 variants. The Australian Government Department of Health [24] aggregates wastewater viral data using flow-adjusted normalization, relying on trend-based classifications rather than absolute thresholds.

To establish a standardized, quantitative aggregation method, the US CDC introduced the WVAL metric as part of the National Wastewater Surveillance System (NWSS). The WVAL method integrates statistical normalization and transformation techniques to ensure comparability between different sewersheds, laboratory methods, and population-adjusted wastewater data. Unlike most pseudo-aggregation approaches, WVAL provides a linear scale, enabling quantitative assessment of viral activity levels at regional and national scales [13].

This study applies the WVAL method to 59 municipal sites in Ontario over a 12-month period, evaluating its effectiveness in aggregating SARS-CoV-2 wastewater data at municipal, regional, and provincial levels. The objective was to assess how well the WVAL metric correlates with clinical case trends and to determine its potential as a standardized wastewater surveillance aggregation tool for public health decision-making.

## Methods

This study utilized wastewater surveillance data collected as part of the Ontario Wastewater Surveillance Initiative (WSI), previously summarized by D’Aoust et al. [10] on behalf of the Ontario WSI Consortium. The dataset comprises 24-hour composite influent samples collected from 109 sites, including 101 wastewater treatment plants (WWTPs) and 8 pumping stations. Six sites also sampled primary sludge. Sample collection followed prescribed field best practices and quality control (QA/QC) protocols, as outlined in the Ontario WSI Protocol [25].

Thirteen academic laboratories and one federal laboratory conducted RT-PCR quantification of viral RNA, targeting N1, N2, E, or N200 [26] gene markers. The pepper mild mottle virus (PMMoV) or daily mean flow rates were used for biomarker-based normalization. Sample frequency varied by site, ranging from daily to weekly analyses. Data were recorded using the Ontario Data Template (ODT) (an MS Excel*^T M^* Workbook) and structured according to the Open Data Model (ODM) for Wastewater-Based Surveillance [27]. Automated QA/QC checks were incorporated into data management before results were compiled in the WSI Ontario Dashboard for visualization and analysis.

To facilitate regional aggregation, we applied the Wastewater Viral Activity Level (WVAL) metric as recommended by the US CDC [13]. The National Wastewater Surveillance System (NWSS) team shared Python scripts with the Ontario Ministry of the Environment, Conservation and Parks (MECP) via personal communication, forming the basis of our computational approach. These scripts were transcribed into R [28] and adapted for Ontario-specific datasets. Validation was performed using US CDC-provided datasets to ensure computational accuracy before application to Ontario data.

The following sections detail the data processing workflow, the correlation analysis between WVAL and clinical case counts (CCC), and the visualization methods used to interpret and communicate aggregation results.

### Data Processing Steps

The daily wastewater viral signal (WVS) in gene copies per milliliter (gc/mL) formed the data set used to determine the WVAL. The key data processing steps included several transformations and calculations to ensure consistency and comparability across sites.

Averaging of daily concentration values was performed using duplicate or triplicate measurements of SARS-CoV-2 gene targets, including N1 and N2, N1 and E, N2 and E, or N2 and N200, depending on the laboratory. The raw gene concentration values (gc/mL) were averaged to determine the daily mean WVS. Similarly, pepper mild mottle virus (PMMoV), which served as a fecal indicator, was also measured and averaged.

Normalization and log transformation were then applied. Biomarker normalization was performed by dividing the daily mean WVS by the PMMoV concentration, while flow-population normalization involved multiplying the daily mean WVS by the flow rate (mL/day) and dividing by the sewershed population. A natural logarithm transformation was applied to these values. If flow data were available, the flow-population normalized values were used; otherwise, biomarker-normalized values were selected.

Outlier handling was conducted using a z-score threshold of 4. The mean and standard deviation (SD) of the normalized values were computed for each sampling location, laboratory, gene target, and normalization method combination. Observations with z-scores greater than 4 were removed to ensure data reliability.

Baseline calculation was performed to establish a reference for WVAL values. The 10th percentile baseline and standard deviation were computed for each site, laboratory, gene target, and normalization method combination. A rolling 12-month window, updated every January 1st and July 1st, was used as the baseline timeframe. A minimum of six weeks of data was required to calculate the baseline and SD. If less than six months of data were available, the baseline and SD were updated weekly until the six-month threshold was reached, after which they remained constant until the next scheduled update on January 1st or July 1st.

Weekly mean and WVAL aggregation were performed by calculating the weekly average of WVAL values while removing missing values and ensuring that only flow-population or biomarker-normalized values were present. Only one laboratory method and gene target combination per site were used, following the CDC’s overlap-handling approach for normalization methods and major laboratory techniques. Aggregation was conducted at the Public Health Unit (PHU), regional, and provincial (Ontario) levels.

A median-based linear aggregated scale was used for final WVAL determination. The WVAL values from each site within a selected region were aggregated using the median or 50th percentile value. The final WVAL values were categorized into minimal (WVAL ≤ 1.5), low (1.5 < WVAL ≤ 3), moderate (3 < WVAL ≤ 4.5), high (4.5 < WVAL ≤ 8), or very high (WVAL > 8) and color-coded for visualization purposes [13].

The key equations corresponding to these data processing steps are presented below.

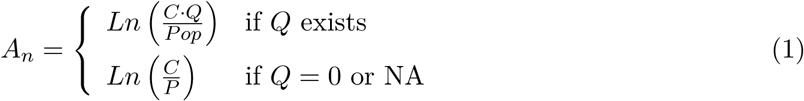

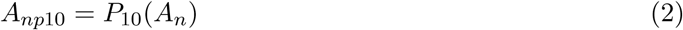

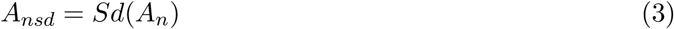

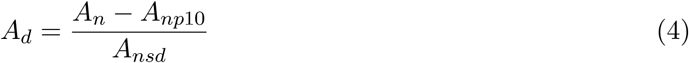

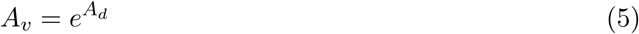

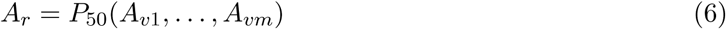

where *A_n_* is the PMMoV (*P*) or Flow (*Q*) normalized natural log-transformed SARS-CoV-2 weekly mean WVS (*C*) in gc/mL over the sampling campaign period for a given sampling location. The flow normalized signal is preferentially selected over the PMMoV if available; the sampling location is characterized by the *P* or *Q* of the sampling location within a region; *A_np_*_10_ is the baseline value defined as the 10-percentile (*P*_10_); *A_nsd_* is the standard deviation for each site; *A_d_* is the number of standard deviations the weekly averaged natural log-transformed concentration values are away from the baseline; *A_v_* is the linear scale converted value that represents the mean WVAL for the week at a sampling location and *A_r_* is the regional median or 50*^th^* percentile of the *A_vi_* values for the group of *m* sampling locations (Sites) (*i* = 1 *…m*) within the aggregation region (*r*) for each week over the total sampling period, respectively.

### Correlation Analysis

To determine how well the WVAL aligned with the COVID-19 clinical case counts (CCC) a correlation analysis was conducted. The total weekly COVID-19 CCC were downloaded from the Public Health Ontario Respiratory Virus Tool. The tool displayed COVID-19 CCC extracted from the Public Health Case and Contact Management Solution (CCM) until June 1, 2024 (extracted on June 4, 2024) [11].

The correlation analysis was performed in two main steps. Data sub-setting was conducted to segment both COVID-19 CCC and SARS-CoV-2 WVAL into distinct epidemiological waves, based on the wave definitions provided by PHO for Waves 1–6 [29]. While the start of Wave 7 was also defined by PHO, its conclusion and the start of Wave 8 were determined through visual inspection of the aggregated wastewater signal [29].

Correlation computation was carried out for each epidemiological wave or season using Spearman’s rank correlation coefficient (*ρ*) to measure the association between WVAL and CCC. The cor.test() function in R [28] was used for statistical computation. Wave 1 was excluded from the analysis due to the limited availability of wastewater surveillance data during its time frame, while Waves 2–8 were included in the correlation assessment.

To visually represent the relationship between WVAL and CCC, scatter plots were generated for each wave, with data points color-coded by date to highlight temporal trends. A linear trend line was added using the geom_smooth() function with the linear model (lm) method to enhance visual interpretation. The statistical and correlation results were summarized in tabular format for further analysis.

### Visualizations of Aggregations

Several visualizations were employed to effectively communicate and interpret the findings. First, the base virus signal data were combined with a shapefile of Ontario public health units (PHUs) [30] to generate static weekly-scaled WVAL maps. Additionally, animated maps were created but are not shown in this study. Second, weekly aggregation bar plots were produced to illustrate time series trends, displaying WVAL results from individual sampling locations, which were then aggregated at the regional and provincial levels. Finally, weekly overlay time series line plots were generated at the site, PHU, and PHO regional levels, enabling direct comparisons of wastewater viral activity trends across different geographic scales.

#### R Packages Used

The manuscript authoring, data analysis and visualizations were conducted using RStudio [31] and the R statistical language [28] on Windows 10 x 64 (build 19045). An important group of R-packages were used which included: FSA [32], ggthemes [33], gridExtra [34], lubridate [35], Hmisc [36], rms [37], doBy [38], neatRanges [39], ggpubr [40], ggside [41], fmesher [42], report [43], tibble [44], rgl [45], RGraphics [46], RColorBrewer [47], gifski [48], sf [49, 50], GGally [51], tmaptools [52], gtools [53], reshape2 [54], plyr [55], ggplot2 [56], forcats [57], stringr [58], tidyverse [59], readxl [60], usethis [61], dplyr [62], purrr [63], readr [64], devtools [65], scales [66], tidyr [67], cowplot [68], bookdown [69], knitr [70] and zoo [71].

## Results and Discussion

This study focused on demonstrating the US CDC WVAL metric aggregation methodology applied to SARS-CoV-2 wastewater surveillance data in Ontario [10]. While this analysis centered on SARS-CoV-2, a similar approach could be extended to other wastewater-based surveillance (WBS) targets, such as influenza A virus. Recently, the US CDC proposed a wastewater influenza A virus level metric, comparing recent WVS data to the 2023–2024 influenza season (October 1, 2023, to March 2, 2024). This metric follows a categorical classification (Minimal to High) and a linear scale using 20-percentile bin thresholds [13].

The WVAL method was applied to 59 municipal sites in Ontario, aggregated at three levels: 34 Public Health Units (PHUs), seven Ontario PHO Regions, and the provincial level. A key assumption in this multi-level aggregation is that the WVAL metric provides a consistent and robust measure of viral activity, ensuring comparability across municipal and regional boundaries through normalization techniques.

Normalization through flow and population adjustments ensures that WVAL remains responsive and comparable across both temporal and spatial scales. Additionally, PMMoV normalization, which accounts for fecal load variations, inherently adjusts for flow dilution and population density differences, further enhancing the consistency of WVAL values. It is assumed that sewershed-specific characteristics, such as mean travel time and hydraulic retention, are inherently managed through the normalization and statistical aggregation process. However, further research is needed to refine this approach by developing a sewershed equivalency statistic, which would assess WVAL-CCC correlations at a sewershed level to account for potential regional variations.

To illustrate the application of WVAL, the Ontario Central East Region (CER) was selected as a detailed case study, with regional summaries for other PHO Regions provided in the Appendix. A correlation analysis was conducted to examine the relationship between clinical case counts (CCC) and WVAL, particularly during major COVID-19 waves. This analysis assessed how well WVAL aligned with CCC trends and explored as a new metric to support public health decision-making.

### Wastewater Signals and Aggreagations

The Central East Region (CER) encompassed three Public Health Units (PHUs) and six municipal sampling sites in central eastern Ontario. The PHUs included Haliburton, Kawartha, and Pine Ridge District Health Unit, Peterborough Public Health, and the Simcoe Muskoka District Health Unit. The individual sampling sites were located in Kawartha Lakes, Cobourg, Barrie, Midland, Peterborough, and Cavan Monaghan.

Figures 1 and 2 present log-scale time-series plots of the observed wastewater viral signals (WVS), measured in gene copies per milliliter (gc/mL), from June 11, 2023, to June 25, 2024. The plots include observed data points, the reported sample limit of detection (SLOD), and a fitted data curve with a 95% confidence envelope. These figures depict raw WVS measurements before normalization, highlighting similarities and differences in general trends across sampling locations.

**Figure 1:**
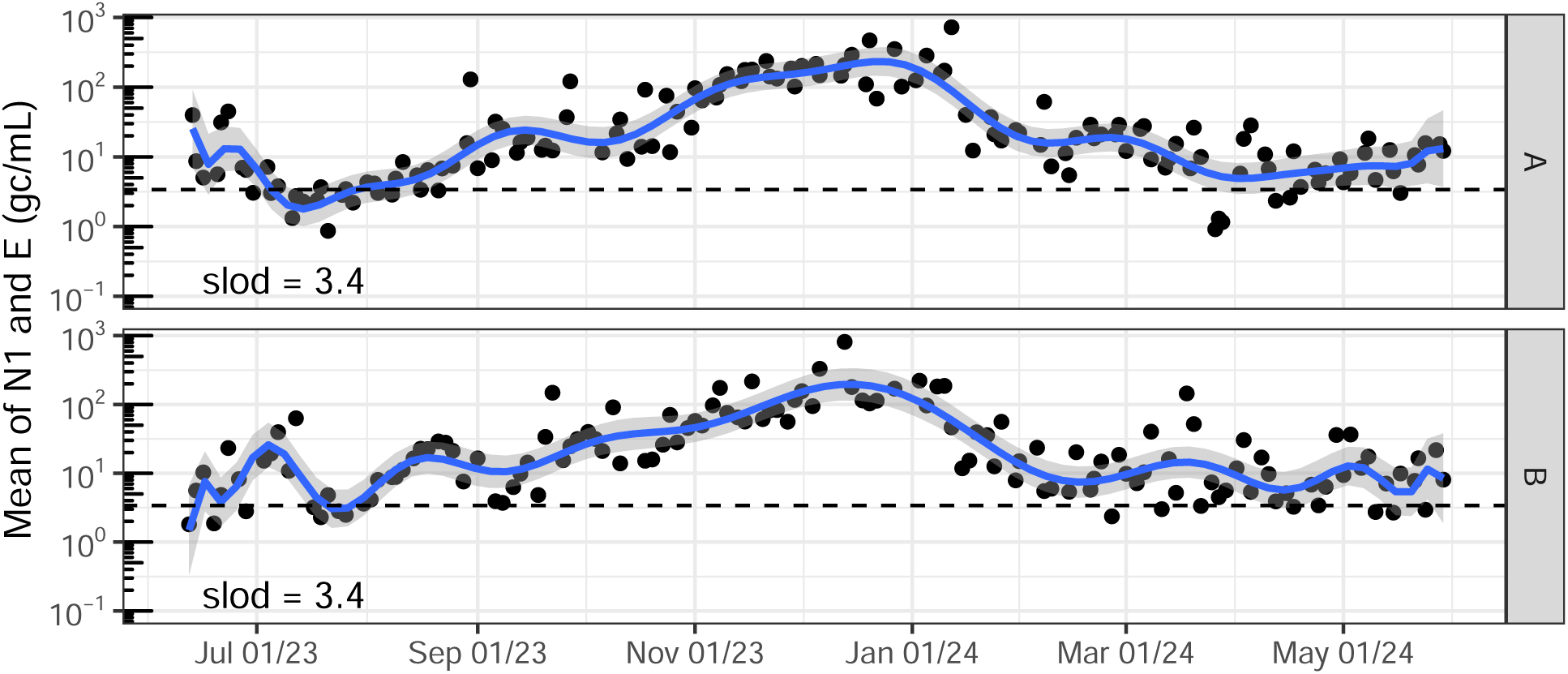
SARS-CoV-2 wastewater signal (WVS) reported mean values with spline curve fit and 95% confidence envelope. A and B refer to Kawartha Lakes and Cobourg, respectively.

**Figure 2:**
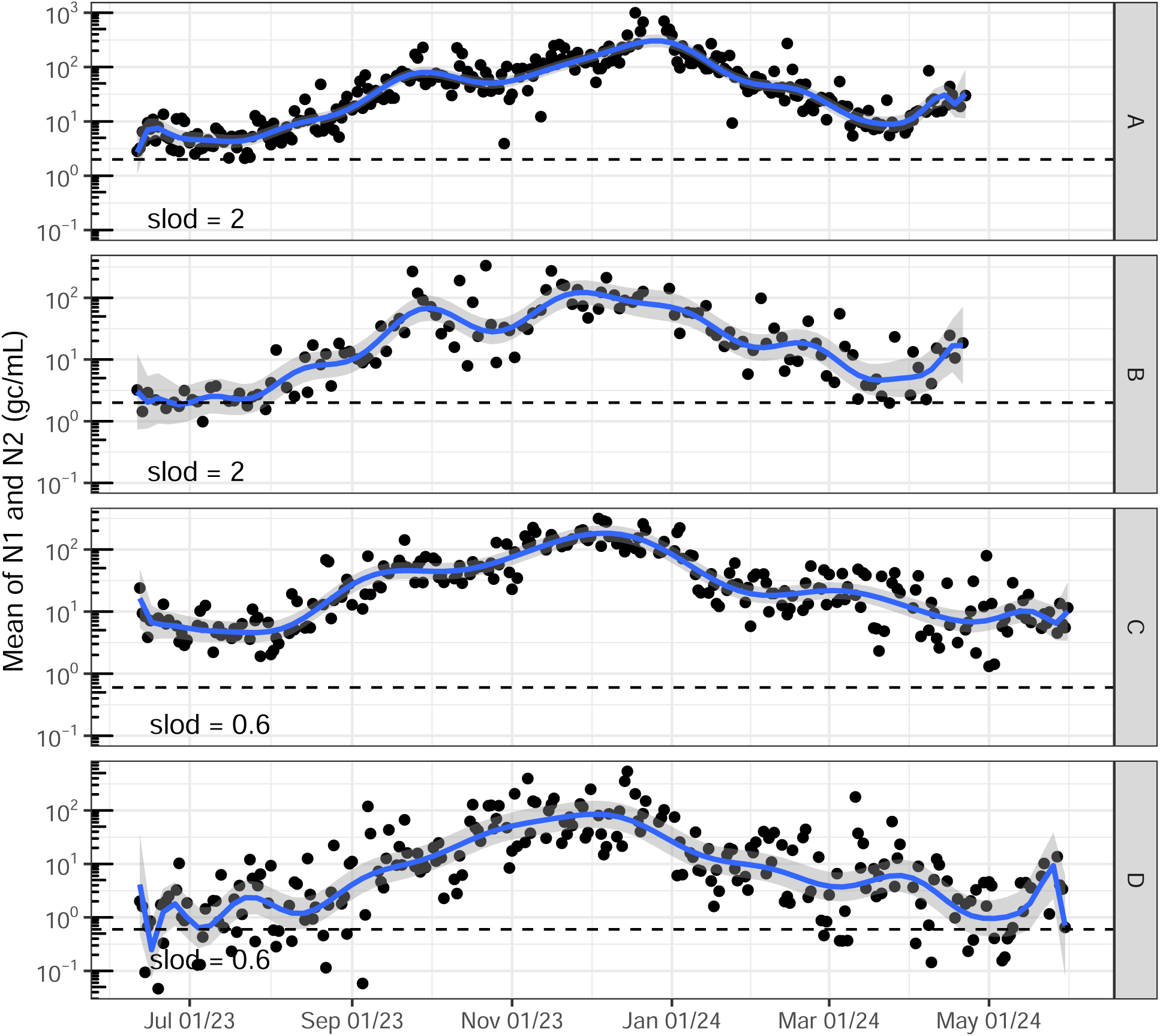
SARS-CoV-2 wastewater signal (WVS) reported mean values with spline curve fit and 95% confidence envelope. A to D refer to Barrie, Midland, Peterborough and Cavan Monaghan, respectively.

Figure 3 presents a time-series bar plot of weekly WVAL for each of the six CER PHO Region sites, displayed individually without aggregation. Weeks with no collected data are indicated by the absence of bars. These site-specific WVAL values served as the basis for higher-level aggregation at the PHU, regional, and provincial scales. The color-coding follows the US CDC classification system.

**Figure 3:**
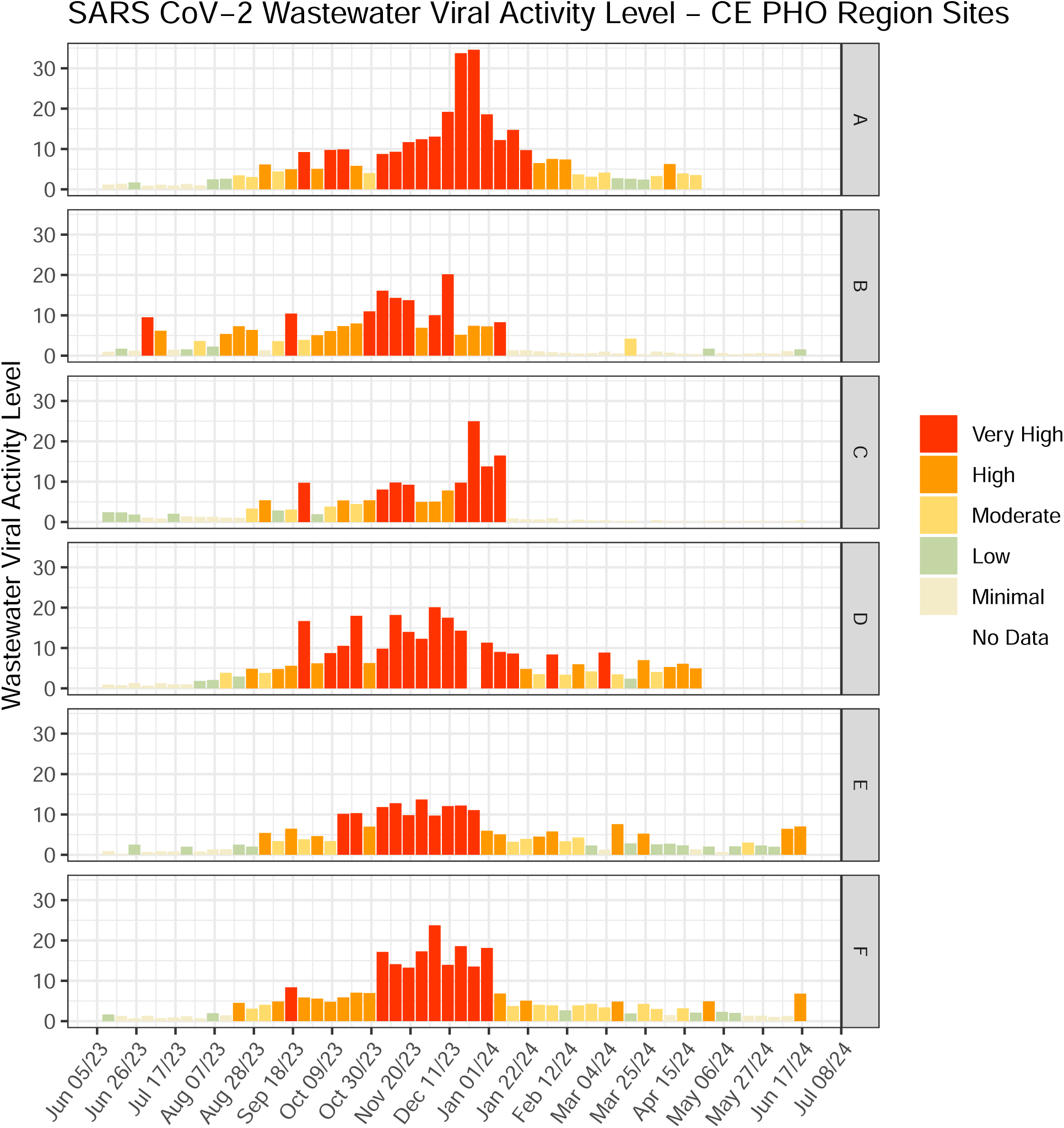
SARS-CoV-2 wastewater viral activity level in the Central East PHO Region sites. A to F refer to Barrie, Cobourg, Kawartha Lakes, Midland, Cavan Monaghan and Peterborough, respectively.

The figure reveals clusters of very high viral activity that fluctuate both spatially and temporally. One potential application of these time-series bar plots is to identify and compare the emergence and progression of COVID-19 surges across locations, providing valuable insights for preemptive public health interventions aimed at mitigating viral spread.

Figure 4 presents an overlay plot illustrating the relative influence of each sampling location within the CER PHO Region. This visualization highlights the contribution of individual sites to the overall regional aggregation. For instance, the City of Barrie (A) exhibited a pronounced peak between December 2023 and January 2024, which significantly impacts the regional aggregation trends observed in Figure 5.

**Figure 4:**
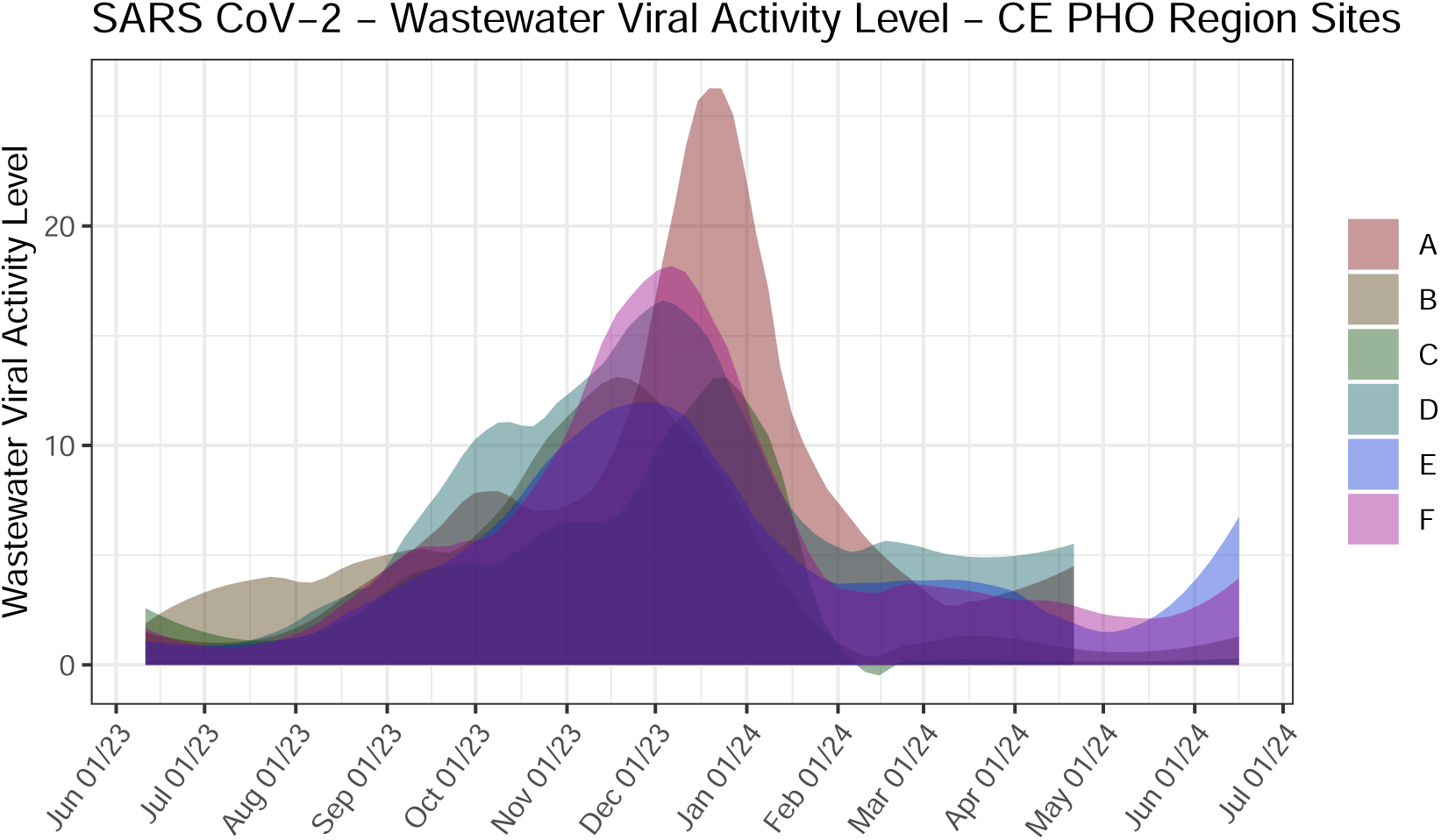
Aggregated SARS-CoV-2 wastewater viral activity level in the Central East PHO Region sampling locations. A to F refer to Barrie, Cobourg, Kawartha Lakes, Midland, Cavan Monaghan and Peterborough, respectively.

**Figure 5:**
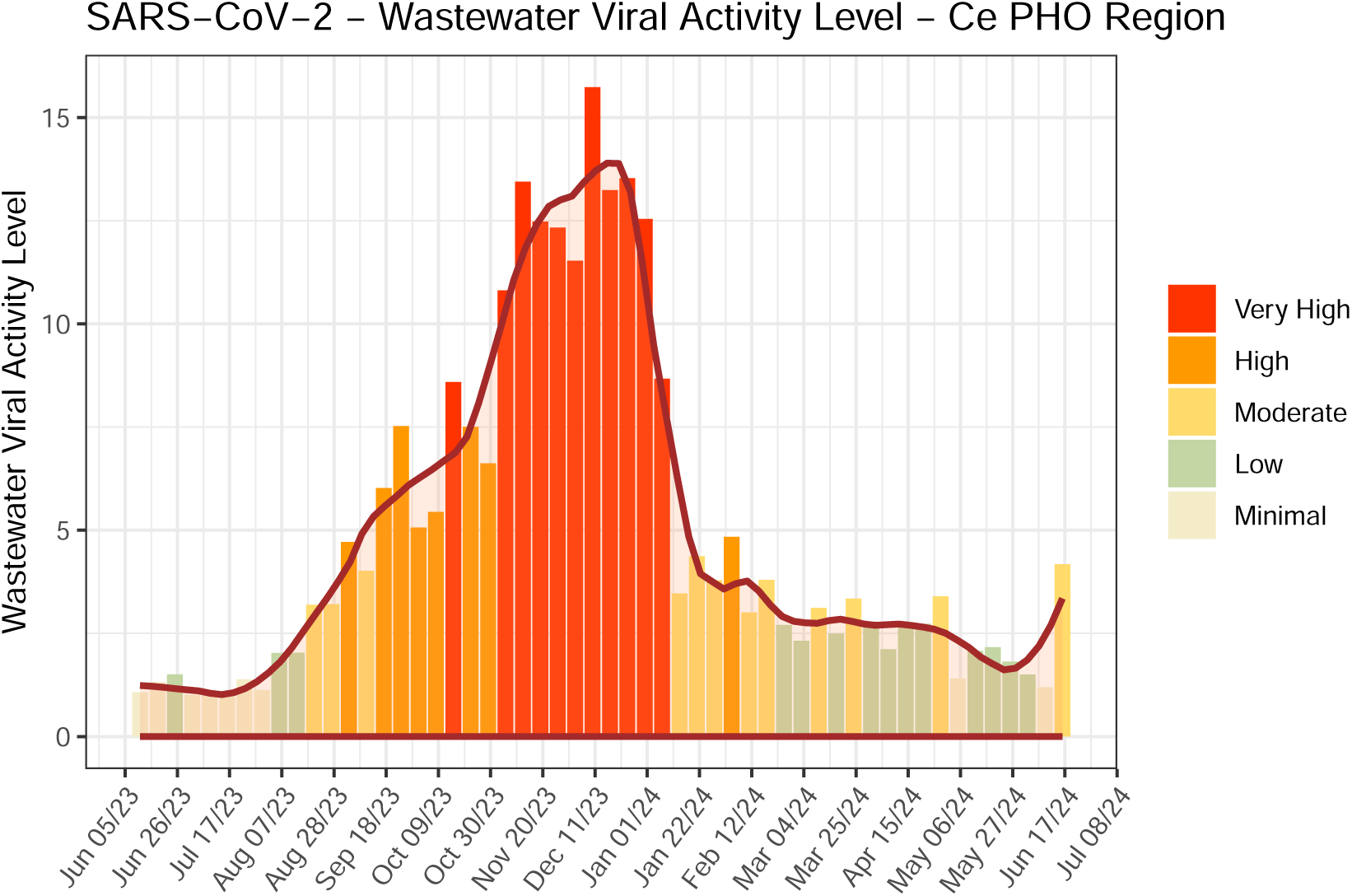
Aggregated SARS-CoV-2 wastewater viral activity level bar and overlay plot for Central East Region with color scale.

Figure 5 displays the regional aggregation bar plot alongside a fitted overlay curve based on the median WVAL values across the six CER sampling sites. The curve fitting was performed using a locally weighted regression (LOWESS) second-order polynomial, with a span parameter of 0.2, allowing for the detection of localized peaks and valleys in WVAL trends. This figure provides a comprehensive regional overview, offering insight into the aggregated wastewater viral activity trends for the CER PHO Region.

Figure 6 presents a time-series WVAL comparison across the three PHUs in the Central East (CE) Region, with threshold levels represented by horizontal lines. The figure highlights how the Simcoe Muskoka District Health Unit experienced a very high WVAL surge earlier than the other two PHUs. Additionally, in the weeks following the major surge, Simcoe Muskoka maintained elevated WVAL levels, remaining consistently higher than the other two PHUs.

**Figure 6:**
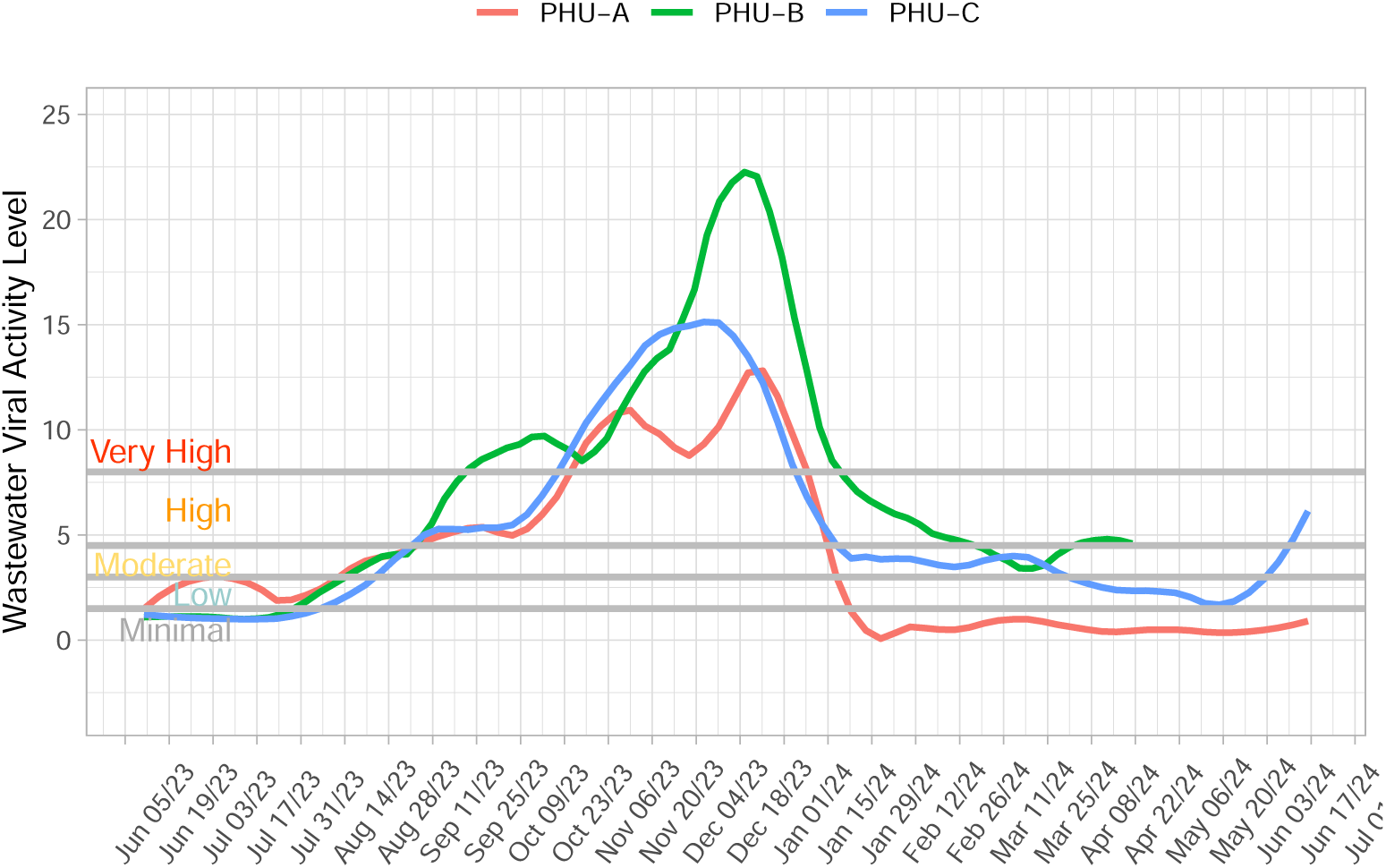
Aggregated SARS-CoV-2 wastewater viral activity level by PHU within Central East Region. PHU-A to PHU-C refer to Haliburton Kawartha Pine Ridge District Health Unit, Simcoe Muskoka District Health Unit and Peterborough Public Health, respectively.

This type of visualization provides a valuable tool for identifying dominant or more heavily impacted PHUs ahead of others. By detecting early surges in specific regions, such plots can aid in anticipating and responding to shifts in community viral activity, supporting targeted public health interventions.

The provincial WVAL aggregations were subsequently generated, with Figure 7 displaying the Ontario weekly WVAL aggregation, fitted using a LOESS curve [56]. Figure 8 illustrates the weekly regional WVAL trends from June 2023 to June 2024. These visualizations allow for a comprehensive assessment of both provincial and regional COVID-19 trends by examining the spatial and temporal distribution of viral activity across Ontario.

**Figure 7:**
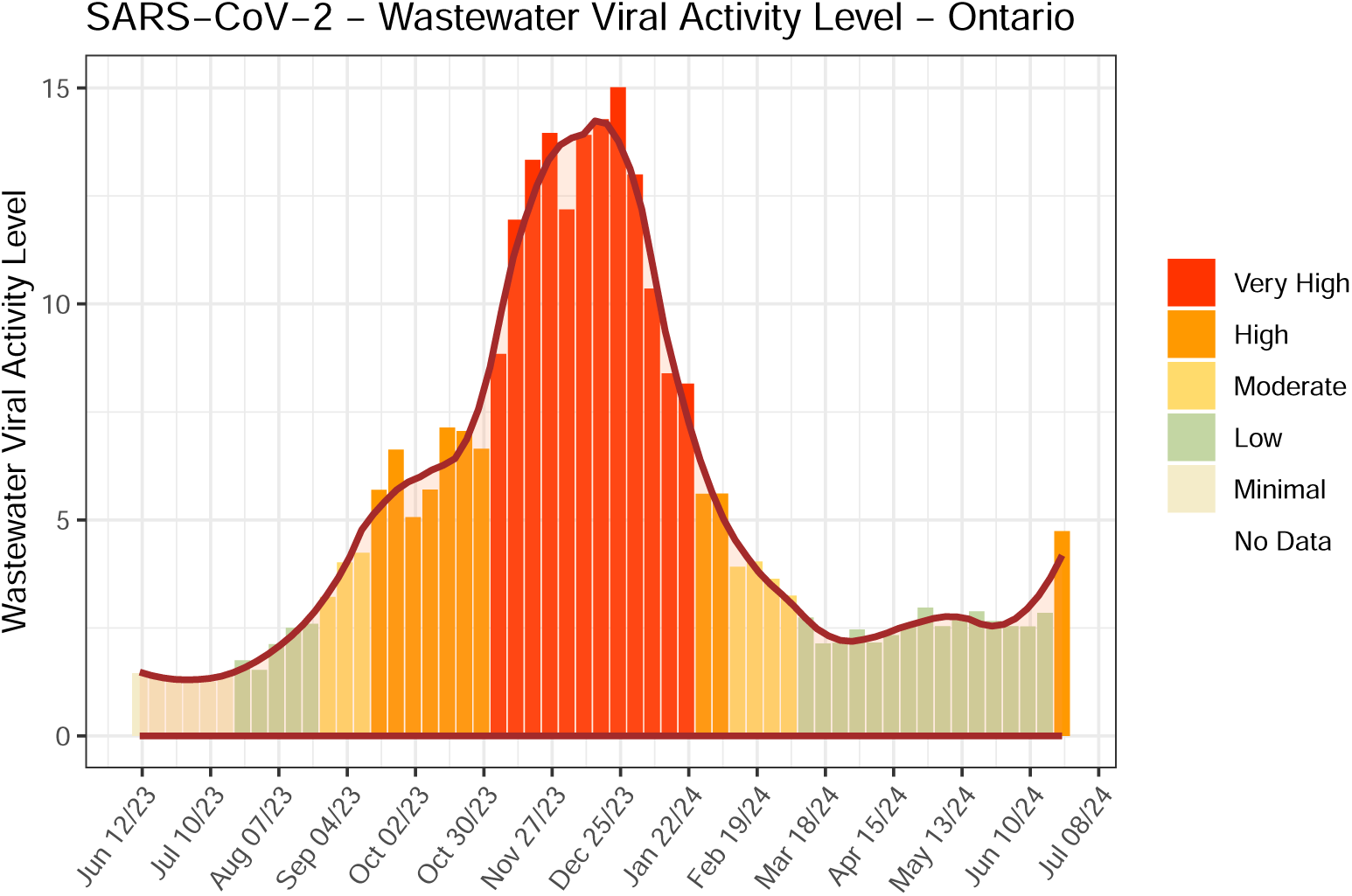
Aggregated SARS-CoV-2 wastewater viral activity level in Ontario with smooth line fit.

**Figure 8:**
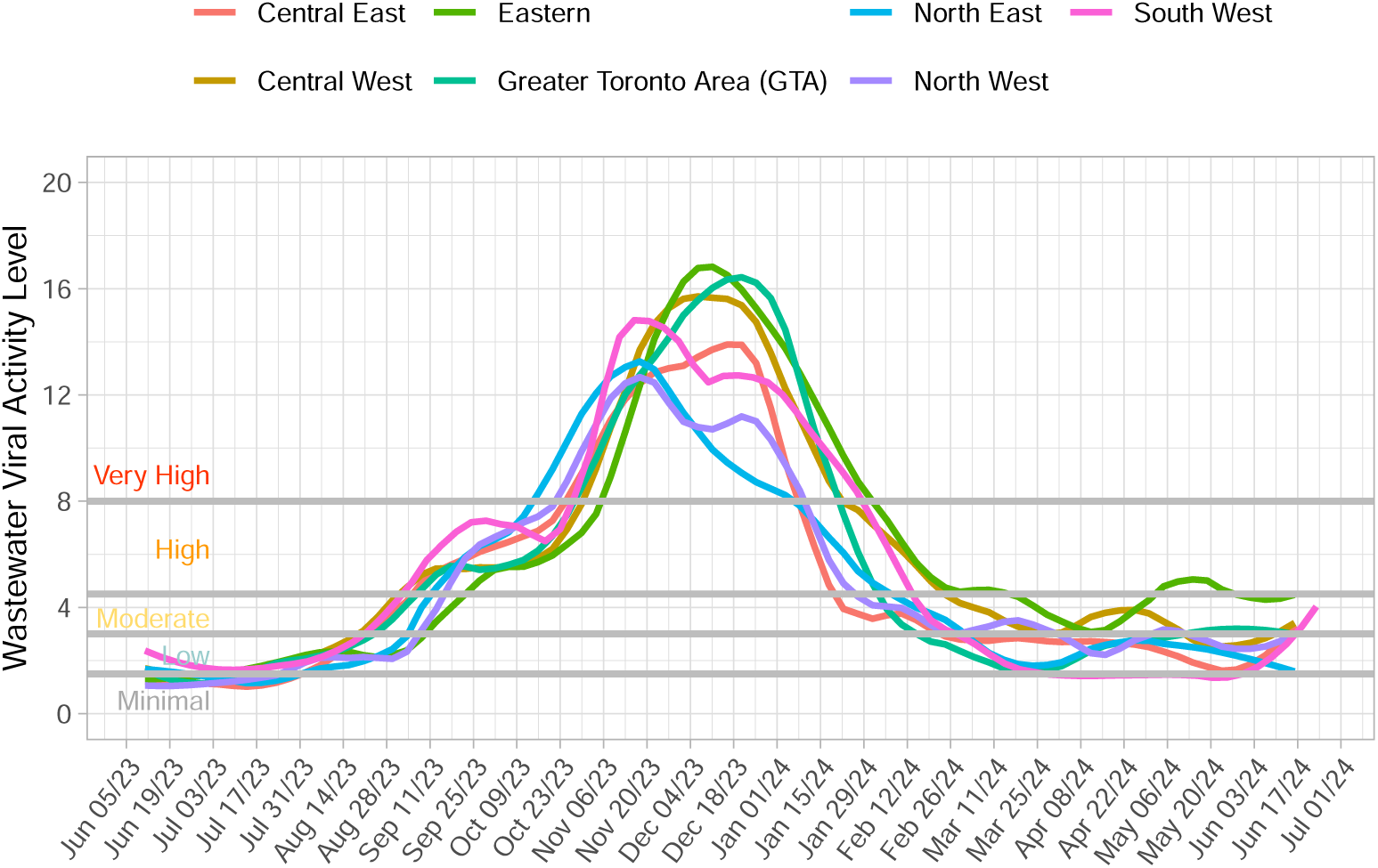
Aggregated SARS-CoV-2 wastewater viral activity level by Region.

From Figure 8, the North East Region exhibited the earliest surge into very high WVAL levels, leading all other regions by one week, followed by the North West Region. The remaining regions entered the very high WVAL category within 1 to 4 weeks of the North East surge. Across all regions, the main WVAL surge lasted approximately 10 to 12 weeks, before declining to high WVAL levels for 2 to 5 weeks. Subsequently, WVAL values transitioned into moderate to low levels, except in the Eastern Region, which experienced a temporary resurgence into high WVAL for three weeks before returning to moderate levels.

These regional WVAL line overlays provide an effective means to compare spatial and temporal trends, offering valuable insights for targeted public health interventions. The remaining six provincial regions were analyzed similarly (not shown) and exhibited comparable trends. A summary of regional aggregations is provided in the Appendix, with Figures 13 to 18 detailing the results for the Northwest, Northeast, Eastern, Greater Toronto Area, South West, and Central West PHO Regions. In all cases, the WVAL metric demonstrated consistent robustness, maintaining a similar scale range across regions.

Figure 9 presents a 12-week area map array depicting the weekly viral activity levels across 34 PHUs during Epiweeks 7–18 (February 11, 2024, to May 4, 2024). This sequential provincial mapping provides a temporal perspective of SARS-CoV-2 activity across Ontario, illustrating how viral trends evolve at the PHU level over time.

**Figure 9:**
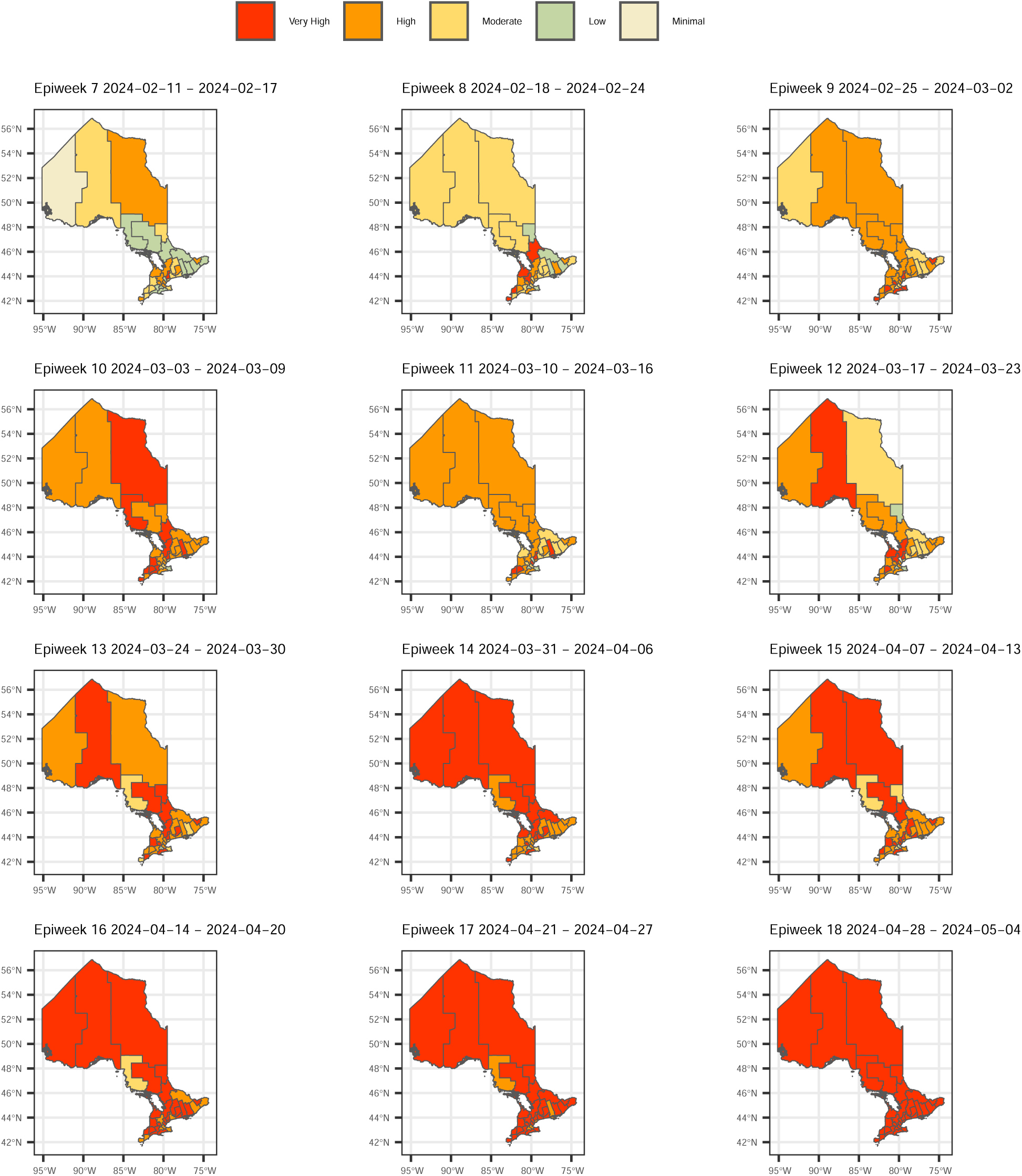
SARS-CoV-2 wastewater viral activity level in Ontario by PHUs for Epiweek 7 - 18, February 11, 2024 to May 4, 2024.

This map-based framework can be further developed to analyze spatial and temporal directional trends in viral spread. Ongoing work aims to incorporate spatial-temporal analytical tools to enhance the interpretation of viral movement patterns across the province. These tools may provide actionable insights to support targeted public health interventions and improve pandemic response strategies.

The provincial overview can also be partitioned into specific regions of interest for more detailed analysis. Figure 10 presents a 12-week area map array, corresponding to the same time frame as Figure 9, but focused on the most populous areas in southern Ontario, with additional coverage of Renfrew County and North Bay Parry Sound District Health Units.

**Figure 10:**
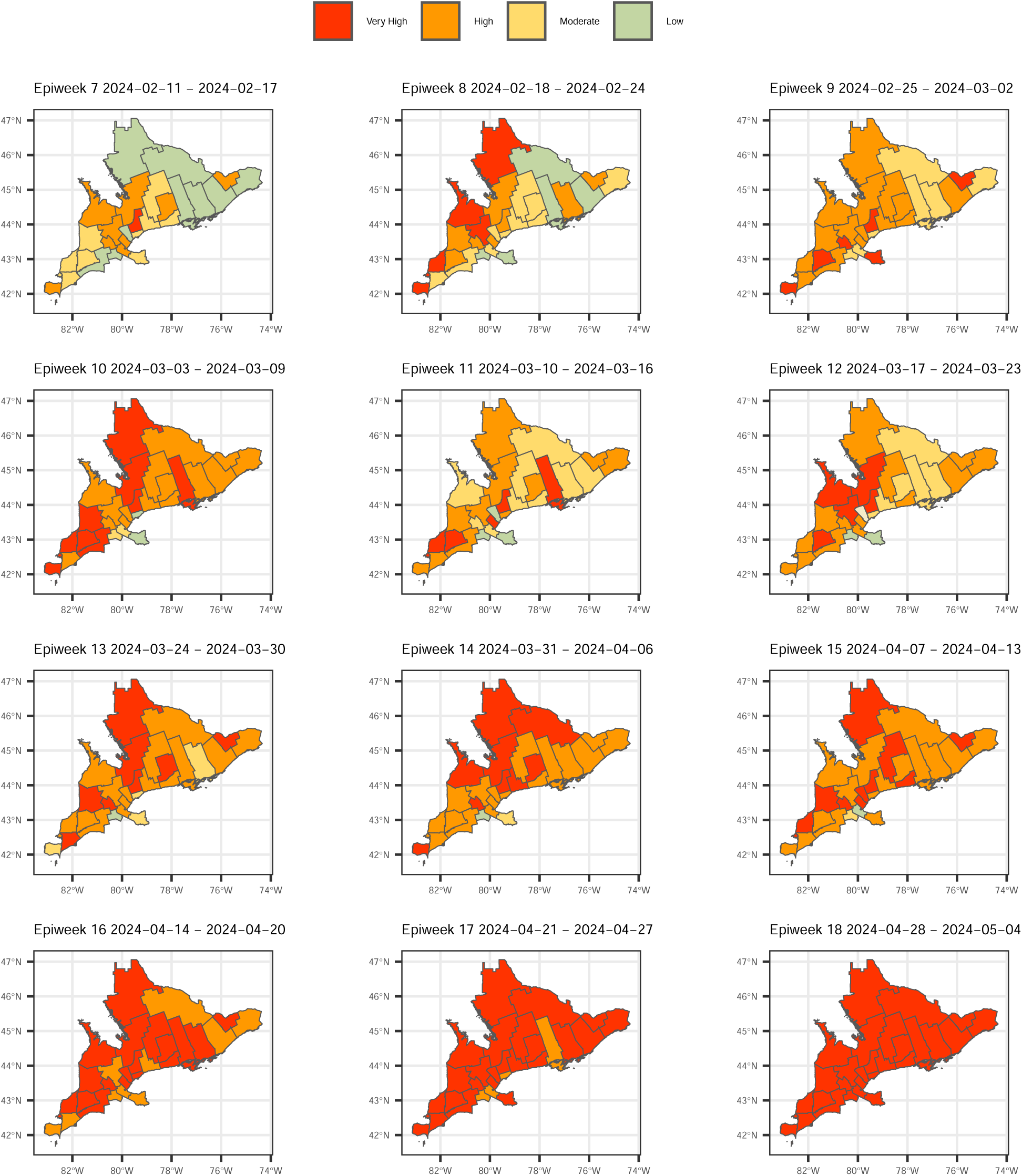
SARS-CoV-2 wastewater viral activity level in Southern Ontario extended with Renfrew County and North Bay Parry Sound District PHUs for Epiweek 7 - 18, February 11, 2024 to May 4, 2024.

This regionalized mapping approach enables a more granular assessment of viral activity trends in high-density areas while still capturing key patterns in less populated but strategically relevant regions. Such analyses can help refine localized public health responses by identifying regional variations in viral spread over time.

### Waves Correlation Analyses

Table 1 and Figure 11 summarize the correlation analysis results, with Table 1 providing the start and end times of each COVID-19 wave, and Figure 11 visualizing the correlation trends. Each wave was analyzed individually to account for shifts in virus transmission dynamics, as well as changes in testing eligibility and test-seeking behavior over time.

**Figure 11:**
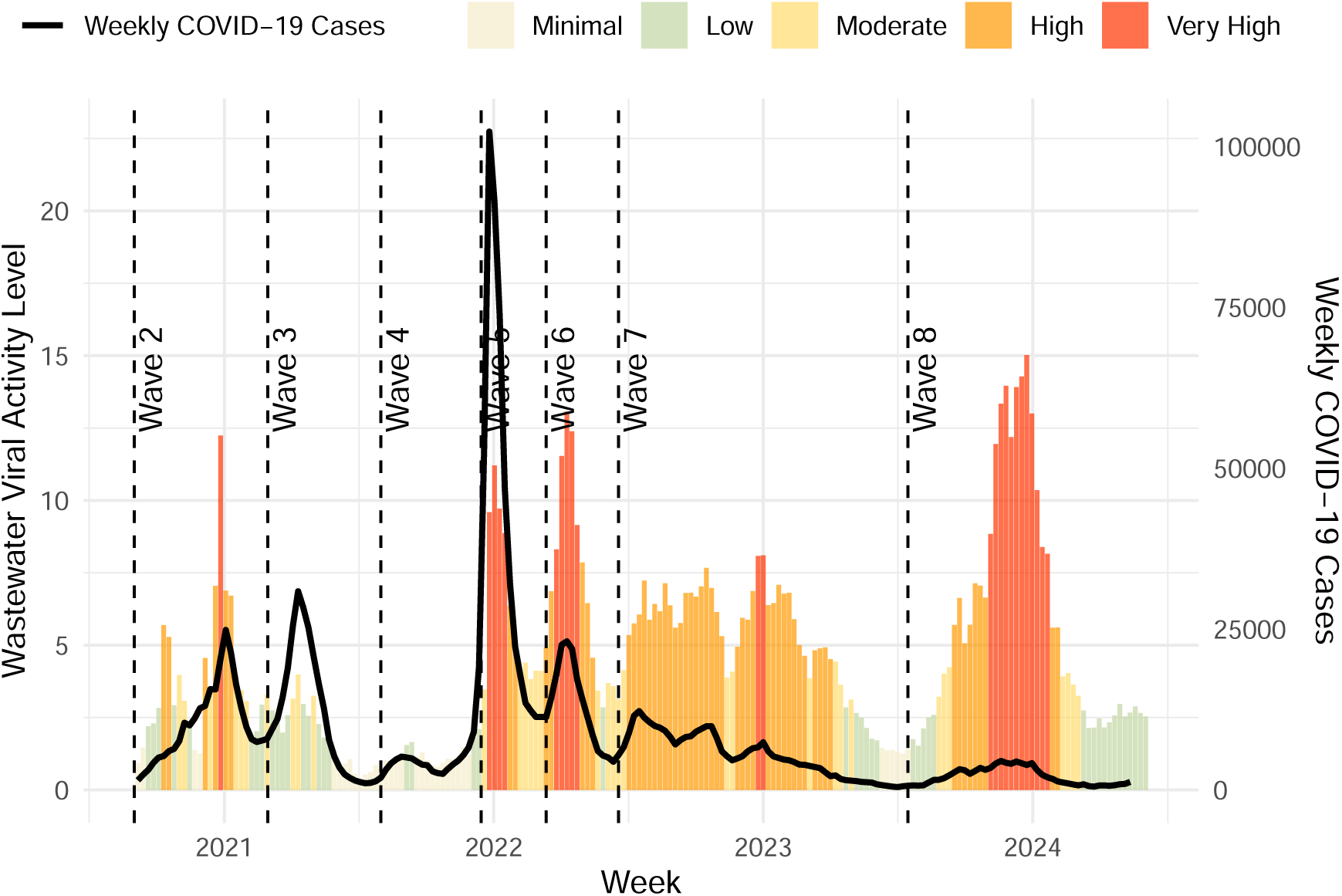
The WVAL and weekly CCC for Wave 2 to 8 from 2021 to 2024

**Table 1:** SARS-CoV-2 WVAL and COVID-19 Clinical Case Counts by Wave.

|  | Wave 2 | Wave 3 | Wave 4 | Wave 5 | Wave 6 | Wave 7 | Wave 8 |
| --- | --- | --- | --- | --- | --- | --- | --- |
| Start Date | 2020-09-01 | 2021-03-01 | 2021-08-01 | 2021-12-15 | 2022-03-13 | 2022-06-19 | 2023-07-16 |
| End Date | 2021-02-28 | 2021-07-31 | 2021-12-14 | 2022-03-12 | 2022-06-18 | 2023-07-15 | 2024-06-03 |
| Spearman's Rho | 0.57 | 0.93 | 0.87 | 0.65 | 0.95 | 0.79 | 0.94 |
| P-Value | <0.05 | <0.05 | <0.05 | <0.05 | <0.05 | <0.05 | <0.05 |
| Maximum WVAL | 12.24 | 3.98 | 2.10 | 11.21 | 13.07 | 8.10 | 15.02 |
| Median WVAL | 3.07 | 1.81 | 1.11 | 4.72 | 6.66 | 5.48 | 4.02 |
| Minimum WVAL | 0.93 | 0.39 | 0.75 | 3.47 | 2.85 | 1.25 | 1.40 |
| Maximum CCC | 24875 | 30883 | 19114 | 102327 | 23106 | 12260 | 4514 |
| Median CCC | 8860.0 | 9499.0 | 4271.5 | 27204.0 | 12724.0 | 5225.5 | 1650.0 |
| Minimum CCC | 1511 | 1044 | 1903 | 11362 | 4386 | 446 | 519 |

Waves 2 to 8 were categorized based on dominant variant types, labeled as Original (Wave 2), Alpha (Wave 3), Delta (Wave 4), Omicron BA.1 (Wave 5), Omicron BA.2 (Wave 6), Omicron BA.5/BQ/XBB.1. (Wave 7), and Omicron EG.5/JN.1 (Wave 8). The corresponding dates for each wave are provided in Table 1 [72–74].

The Spearman correlation values were all statistically significant at the 95% confidence level (p-value of *<* 0.05) and the *ρ* values were *>* 0.65 for all waves except Waves 2 and 5. The lower correlation (*ρ* = 0.65) in Wave 5 was likely due to the change in case testing eligibility that occurred starting in December 31, 2021, partially through this Wave, with the emergence of the Omicron variant [75]. As well, the lower correlation in Wave 2 (*ρ* = 0.57) is likely due to progressive increases to clinical case testing capacity with the expansion of targeted asymptomatic testing at pharmacies across the province there was an increase in clinical cases detected as a percentage of the number of people actually infected. This is evident in the Wave 2 correlation plot with the red dots above the best fit linear regression line [76–78].

Each scatter plot in Figure 12 represents a different epidemic wave, with WVAL on the x-axis and clinical case counts (CCC) on the y-axis. The color gradient for each point indicates the progression of weeks from blue (early weeks) to red (later weeks) within each Wave. The black line denotes the linear regression fit and the Spearman’s rank correlation coefficient (*ρ*) is provided in Table 1. Overall, the Spearman’s (*ρ*) values indicate a good to excellent agreement between the WVAL and CCC which suggests that the WVAL metric provides a good surrogate measure and predictive tool that aligns well with potential for clinical cases when testing is not available.

**Figure 12:**
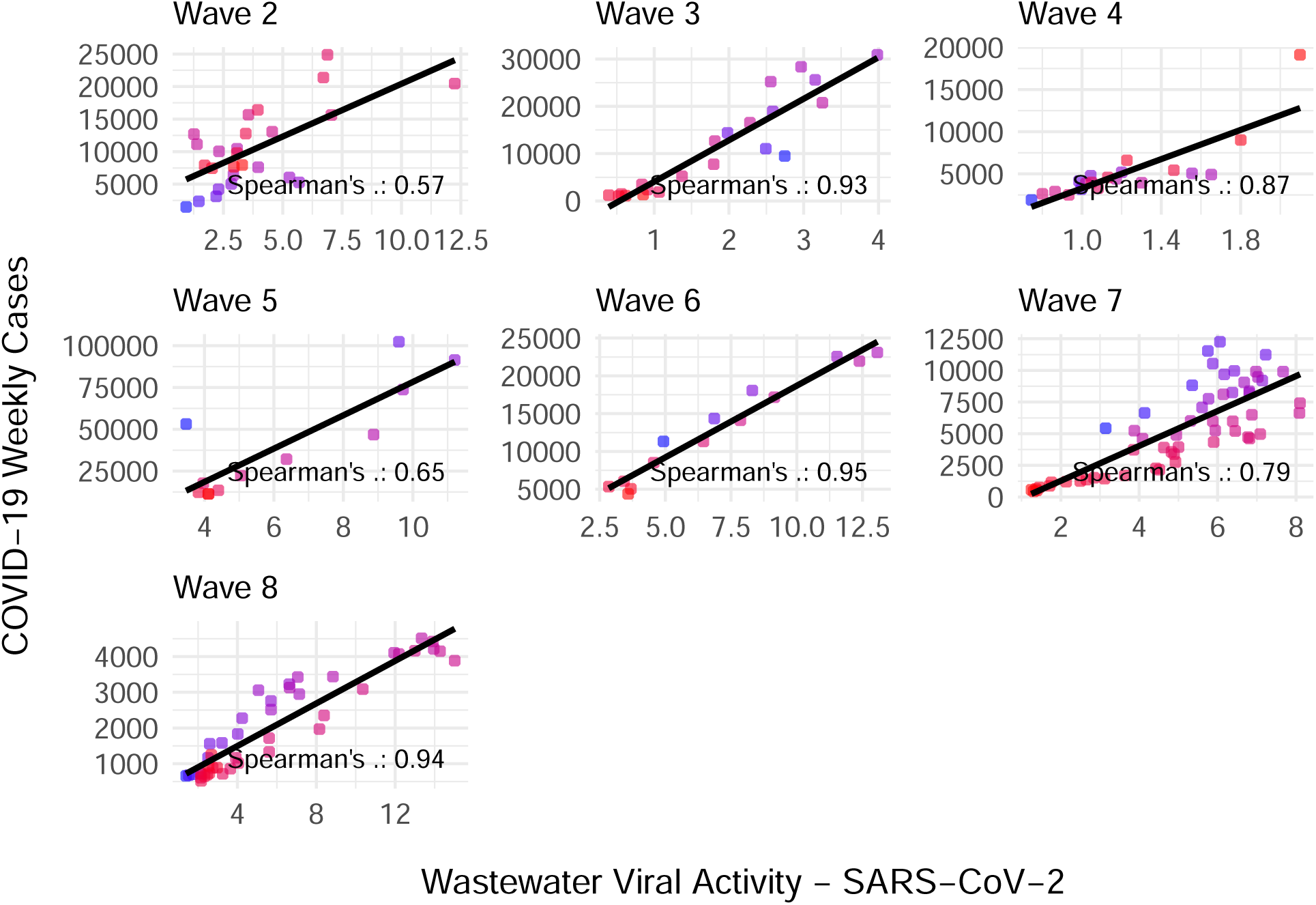
Scatter plots of SARS-CoV-2 WVAL versus clinical case counts by Wave and linear regression fit.

**Figure 13:**
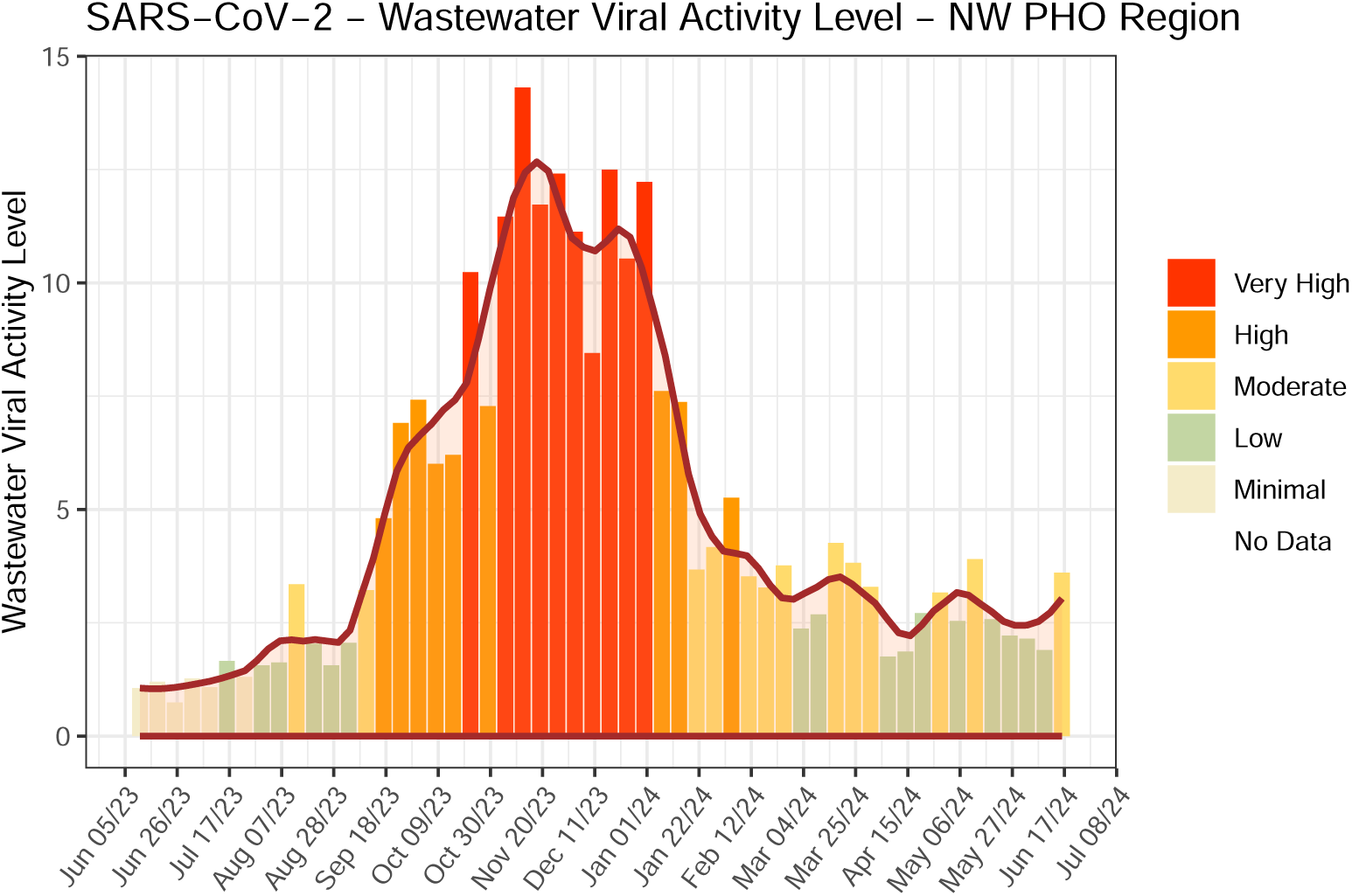
Northwest (NW) PHO Region aggregated SARS-CoV-2 wastewater viral activity level bar and overlay curve fit with color scale.

**Figure 14:**
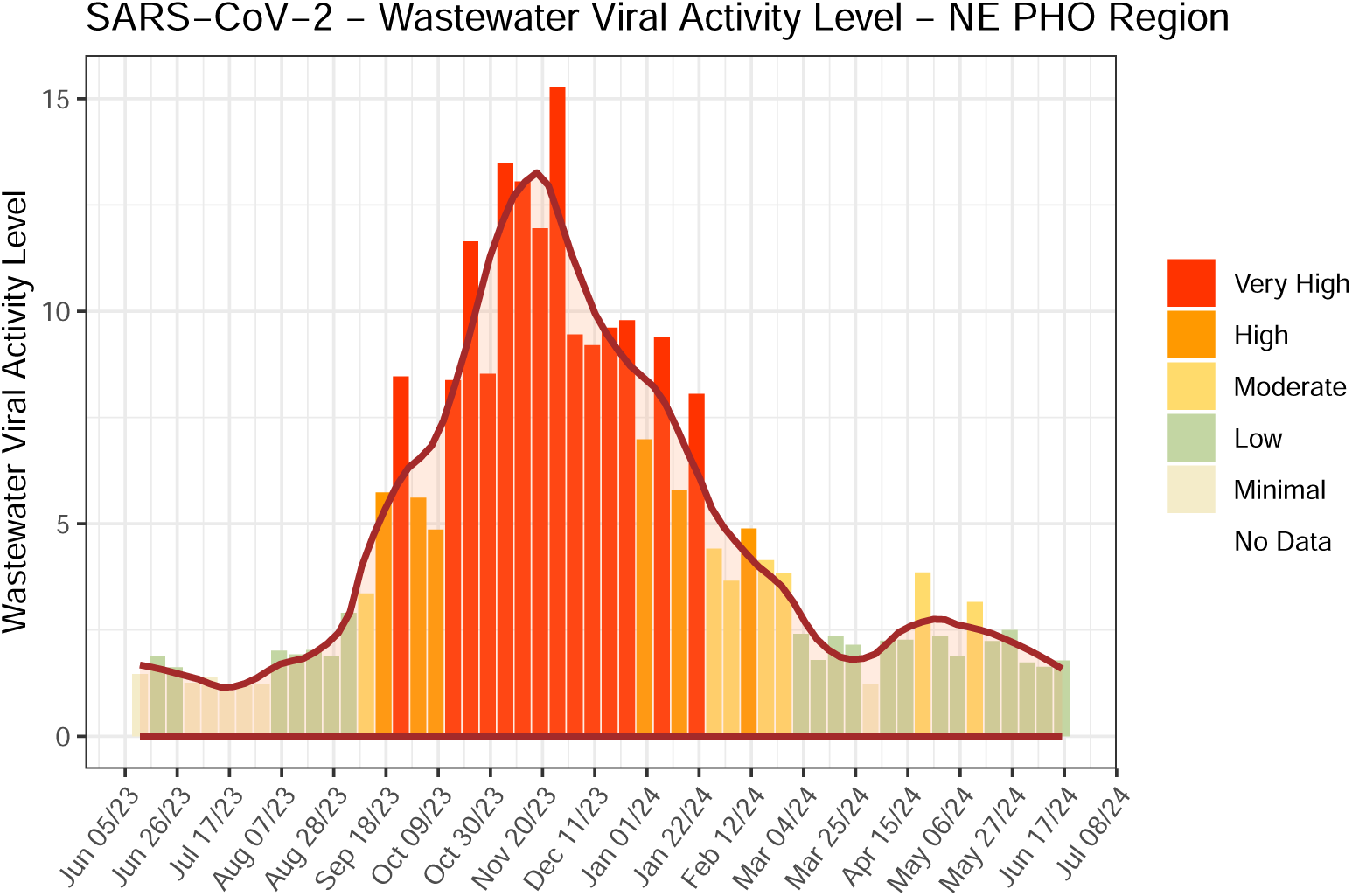
Northeast (NE) PHO Region aggregated SARS-CoV-2 wastewater viral activity level bar and overlay curve fit with color scale.

**Figure 15:**
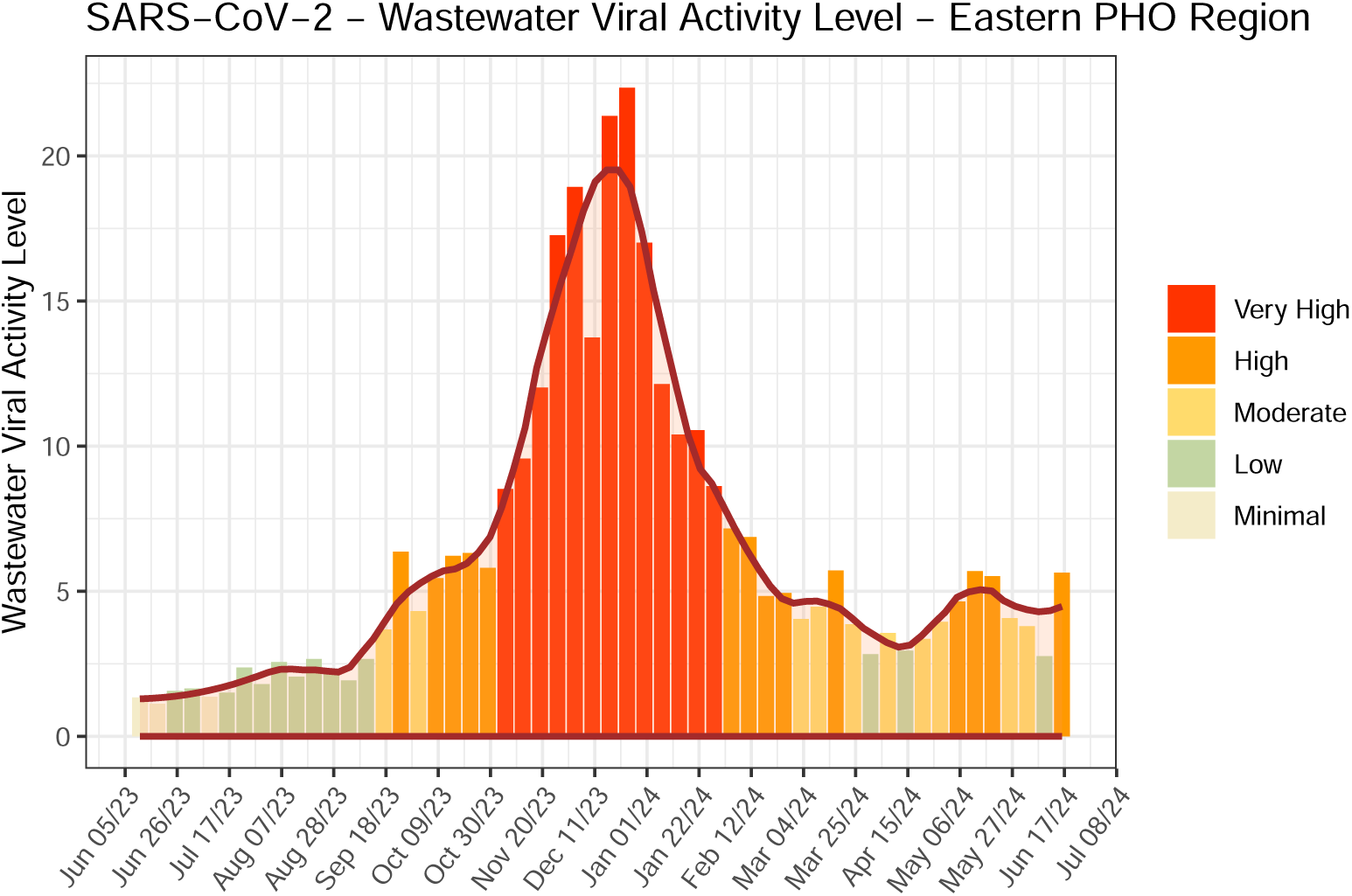
Eastern PHO Region aggregated SARS-CoV-2 wastewater viral activity level bar and overlay curve fit with color scale.

**Figure 16:**
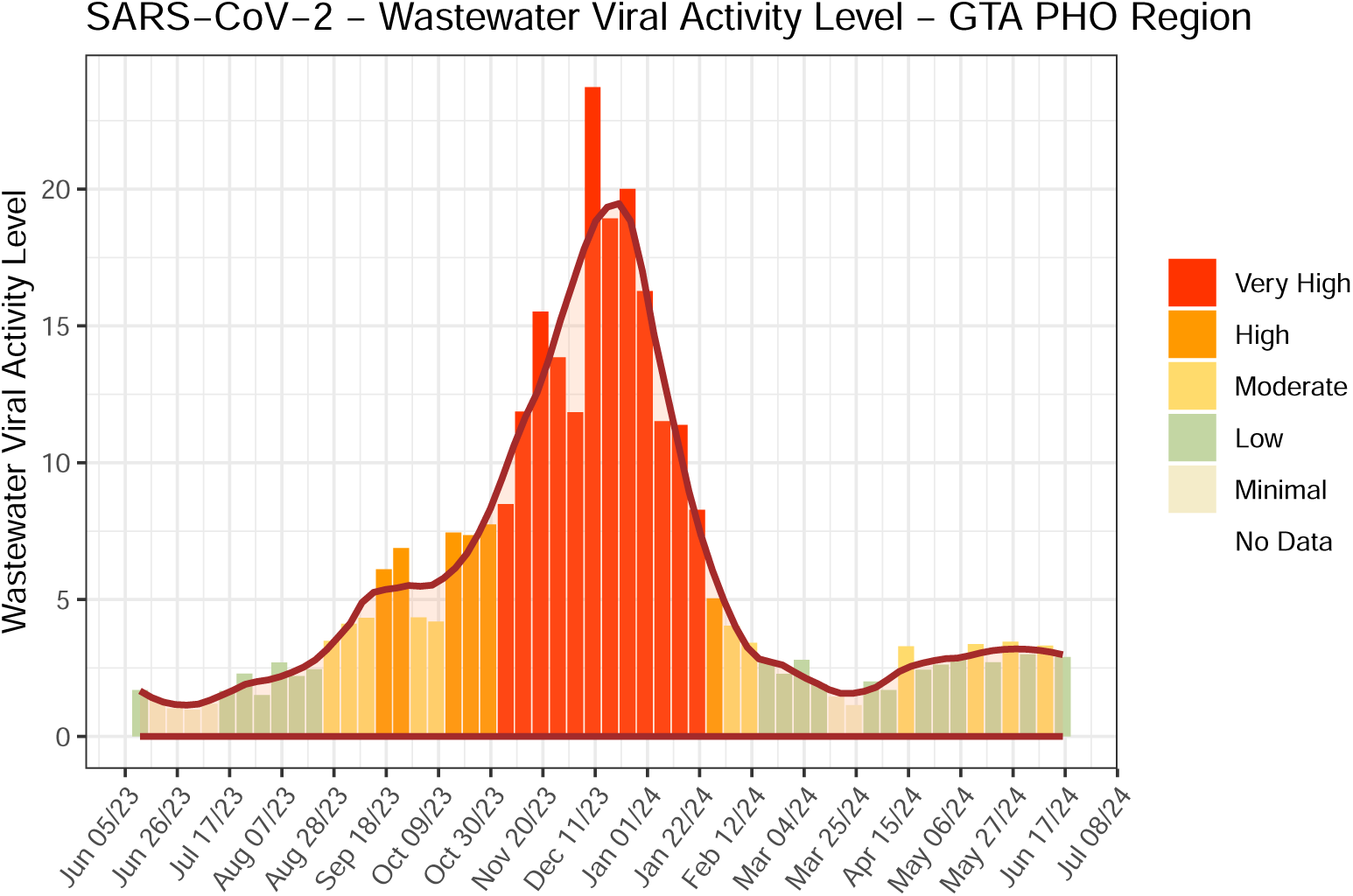
GTA PHO Region aggregated SARS-CoV-2 wastewater viral activity level bar and overlay curve fit with color scale.

**Figure 17:**
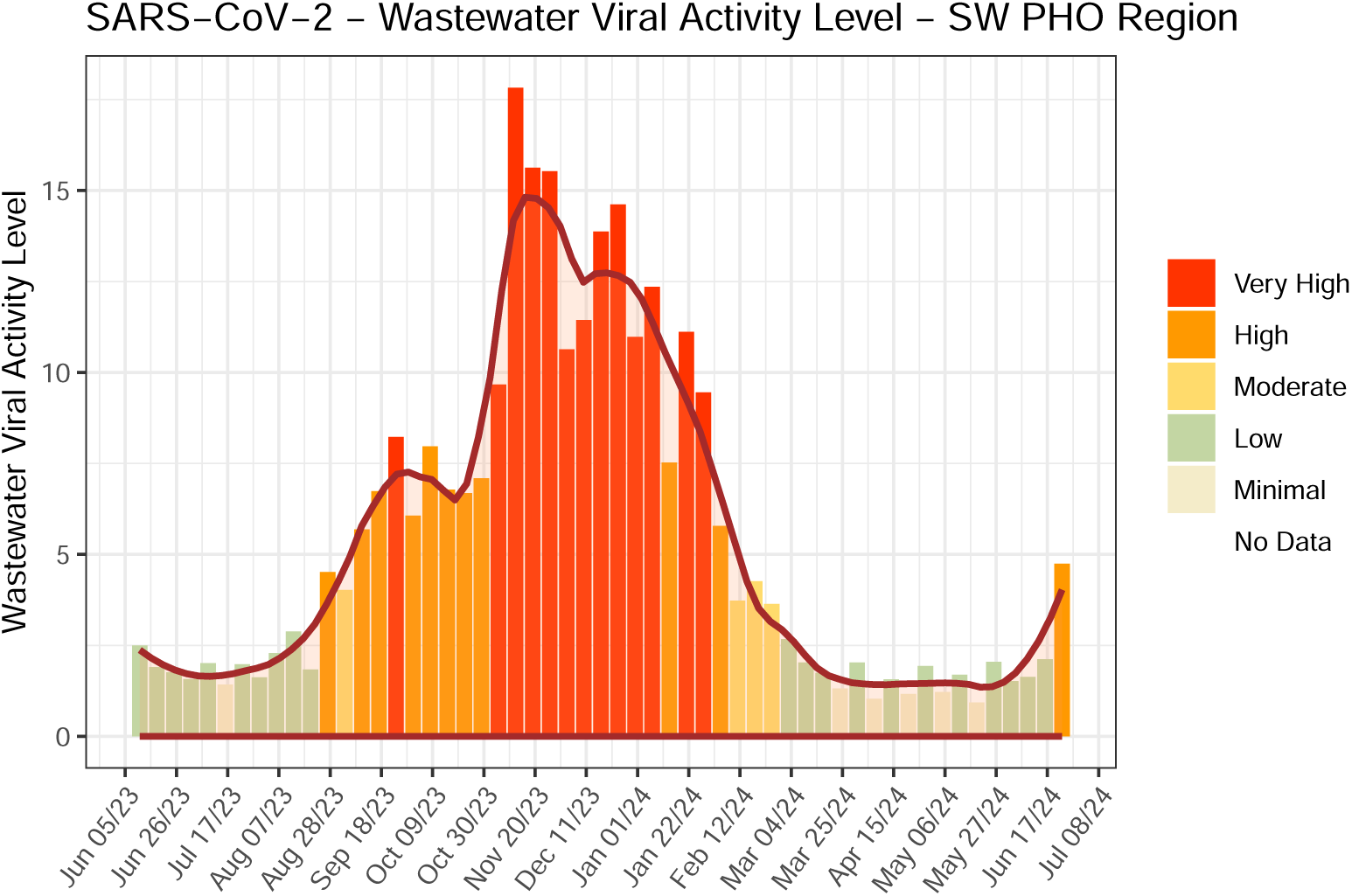
South West (SW) PHO Region aggregated SARS-CoV-2 wastewater viral activity level bar and overlay curve fit with color scale.

**Figure 18:**
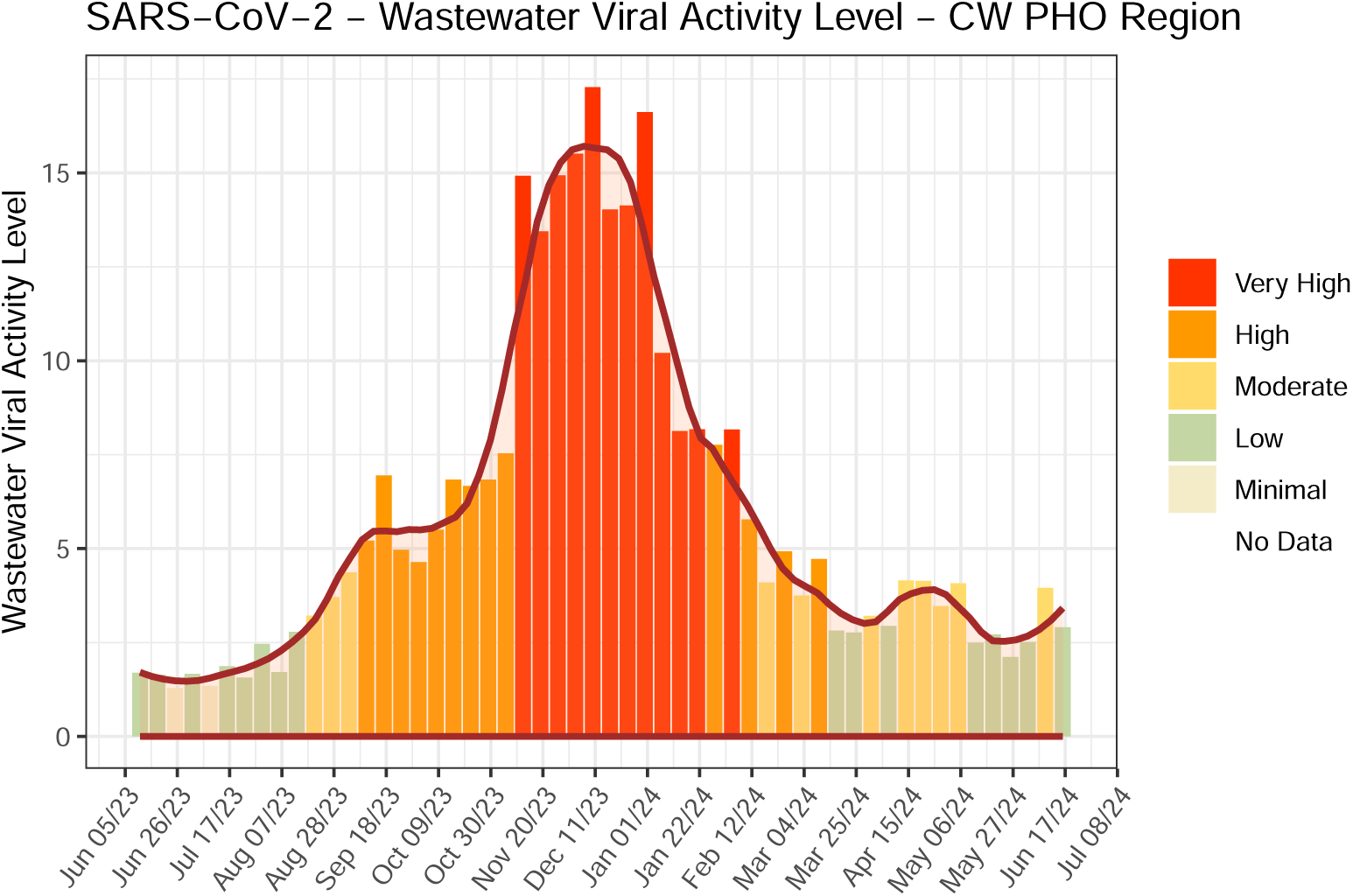
Central West (CW) PHO Region aggregated SARS-CoV-2 wastewater viral activity level bar and overlay curve fit with color scale.

The Spearman correlation values were all statistically significant at the 95% confidence level (p-value *<* 0.05), with *ρ* values > 0.65 for all waves except Wave 5. The lower correlation observed in Wave 5 (*ρ* = 0.57) coincided with the emergence of the Omicron variant [75] and was likely influenced by the dramatic reduction in case testing eligibility across Ontario, which took effect on December 31, 2021, midway through this wave [75].

Each scatter plot in Figure 12 represents a different epidemic wave, with WVAL on the x-axis and clinical case counts (CCC) on the y-axis. The color gradient of the points reflects the temporal progression within each wave, transitioning from blue (early weeks) to red (later weeks). A black regression line indicates the linear trend, while the Spearman’s rank correlation coefficient (*ρ*) for each wave is provided in Table 1.

Overall, the Spearman’s (*ρ*) values demonstrate good to excellent agreement between WVAL and CCC, indicating that WVAL serves as a strong surrogate measure for clinical case trends. This reinforces WVAL’s utility as a confirmatory tool, particularly in situations where clinical testing data is unavailable or limited, providing an alternative means of assessing community infection trends.

### Conclusions and Recommendations

The US CDC WVAL metric and aggregation algorithm demonstrated numerical stability, effectively managing variability in reported data and limits of detection across different sampling locations and laboratories. When analyzed by specific epidemiological waves, the WVAL metric correlated well with clinical case counts (CCC), confirming its robustness as a comparative and confirmatory tool for wastewater-based epidemiology (WBE).

Several key visualization methods were developed to enhance the interpretation of WVAL trends and included time-series weekly bar plots at the PHU, regional, and provincial levels, enabling identification of temporal origins of COVID-19 surges and supporting time-sensitive interventions. Overlay area plots illustrating the relative contributions of each sampling location, aiding in targeting highly impacted areas. Time-series weekly linear overlay plots at the PHU and regional levels, facilitating comparative temporal-spatial assessments to determine both the timing and location of surges across the province. Provincial PHU-based area maps, presenting a time-series of static images by Epiweek, allowing for visualization of spatial and temporal changes in WVAL at the PHU level.

Further statistical and spatial-temporal analysis is under consideration to expand the utility of WVAL, incorporating directional statistical visualizations to better characterize viral movement patterns over time and space.

The WVAL metric could be integrated into PHU-level interpretations of regional health trends by combining wastewater, clinical, and hospitalization data in a weighted analysis. A similar approach is currently employed by Ottawa Public Health, where an overall indicator of respiratory virus activity is calculated using weighted contributions from wastewater viral loads (40%), new hospitalizations (20%), and healthcare institution outbreaks (20%) [79].

An extension of the WVAL metric is being explored to account for differences in wastewater signal responses across sewersheds. This method applies linear regression between CCC and WVS to establish a sewershed equivalency factor (EQF), which adjusts WVAL values to enhance comparability between locations. The EQF is defined as the ratio of a site’s best-fit linear regression curve to that of a reference site, where the reference sewershed is assigned an EQF of 1 and other sites receive values between zero and one. This approach allows for finer regional adjustments, improving the interpretability of WVAL data across diverse sewersheds.

A key challenge associated with aggregation methods is the modifiable areal unit problem (MAUP), which can introduce bias in results based on the chosen aggregation scale. In this study, three different scales were assessed: Local municipal sewershed level (low scale); Public health unit level (mid-scale); Regional scale (large scale) and Provincial level (very large scale).

As aggregation scales increase, local variations may become masked, and statistical measures such as mean, standard deviation, and regression trends can be influenced by the boundaries of aggregation regions [80]. These factors must be considered when interpreting regional and provincial trends.

The validity of any aggregation method in wastewater-based epidemiology depends on the assumption that sewershed characteristics—such as travel time, industrial and stormwater contributions, sedimentation and resuspension, and viral decay—have a minimal impact on the normalized SARS-CoV-2 wastewater viral signal (WVS) between sewersheds, relative to inherent variability within each system. Recent research on sewer transport conditions and the decay of enveloped viruses like SARS-CoV-2 supports the use of PMMoV normalization to correct for time decay effects, particularly when the virus interacts with sewer bed and near-bed conditions [81, 82].

## Appendix

This appendix includes the additional six provincial regional aggregations in Figures 13 to 18 for the Northwest, Northeast, Eastern, Greater Toronto Area, South West and Central West PHO Region, respectively. In all cases the WVAL metric provided similarly robust results within a consistent scale range.

## Data and Code Availability Statement

The data and R-code used in this study are available or accessible on request from the corresponding authors. Due to institutional or privacy restrictions, some datasets may not be publicly shared.

## Data Availability

All data produced in the present study are available upon reasonable request to the authors

https://doi.org/10.1038/s41597-024-03414-w

## Acknowledgments

The authors thank and extend their appreciation to: The Ontario Wastewater Surveillance Consortium for data accessibility and particular all laboratory staff that analyzed the wastewater samples; all the municipal staff, including field staff, that collected samples and that contributed to the wastewater field sampling and Ontario MECP Wastewater Surveillance Initiative lead by Steven Carrasco, Bahar Aminvaziri and Susanne Edwards; the US CDC NWSS Team including Amy Kirby, Heather Reese, Dennis Nichols and Daniel Cornforth. Without the hard work and dedication of this multidisciplinary group of people this publication would not have been possible.

## References

[1] D. G. Manuel, R. Delatolla, D. N. Fisman, M. Fuzzen, T. Graber, G. M. Katz, J. Kim, C. Landgraff, A. MacKenzie, A. Maltsev, A. Majury, R. M. McKay, J. Minnery, M. Servos, J. S. Weese, A. McGeer, K. B. Born, K. Barrett, B. Schwartz, P. Jüni, The role of wastewater testing for sars-cov-2 surveillance, Science Briefs of Ontario COVID-19 Science Advisory Table 2 (40) (2021). doi:10.47326/ocsat.2021.02.40.1.0. URL 10.47326/ocsat.2021.02.40.1.0

[2] G. Chavarria-Miró, E. Anfruns-Estrada, A. Martínez-Velázquez, M. Vázquez-Portero, S. Guix, M. Paraira, B. Galofré, G. Sánchez, R. M. Pintó, A. Bosch, Time evolution of severe acute respiratory syndrome coronavirus 2 (sars-cov-2) in wastewater during the first pandemic wave of covid-19 in the metropolitan area of barcelona, spain, Applied and Environmental Microbiology 87 (7) (2021) e02750–20. doi:10.1128/AEM.02750-20.

[3] G. Bonanno Ferraro, C. Veneri, P. Mancini, M. Iaconelli, E. Suffredini, L. Bonadonna, L. Lucentini, A. Bowo-Ngandji, C. Kengne-Nde, D. S. Mbaga, G. Mahamat, H. R. Tazokong, J. T. Ebogo-Belobo, R. Njouom, S. Kenmoe, G. La Rosa, A state-of-the-art scoping review on sars-cov-2 in sewage focusing on the potential of wastewater surveillance for the monitoring of the covid-19 pandemic, Food and Environmental Virology 14 (4) (2022) 315–354. doi:10.1007/s12560-021-09498-6.

[4] M. J. Joung, C. S. Mangat, E. M. Mejia, A. Nagasawa, A. Nichani, C. Perez-Iratxeta, S. W. Peterson, D. Champredon, Coupling wastewater-based epidemiological surveillance and modelling of sars-cov-2/covid-19: Practical applications at the public health agency of canada, Canada Communicable Disease Report 49 (5) (2023) 166–174. doi:10.14745/ccdr.v49i05a01. URL 10.14745/ccdr.v49i05a01

[5] S. Akingbola, R. Fernandes, S. Borden, K. Gilbride, C. Oswald, S. Straus, A. Tehrani, J. Thomas, R. Stuart, Early identification of a covid-19 outbreak detected by wastewater surveillance at a large homeless shelter in toronto, ontario, Canadian Journal of Public Health 114 (1) (2023) 72–79. doi:10.17269/s41997-022-00696-8. URL 10.17269/s41997-022-00696-8

[6] T. Piggott, M. Kharbouch, M. E. Donaldson, C. Pigeau, D. Churipuy, G. Pacey, C. J. Kyle, Wastewater surveillance for earlier detection of seniors congregate living covid-19 outbreaks in peterborough, ontario, Canada Communicable Disease Report 49 (2/3) (2023) 35–43. doi:10.14745/ccdr.v49i23a02. URL 10.14745/ccdr.v49i23a02

[7] C. Chen, Y. Wang, G. Kaur, A. Adiga, B. Espinoza, S. Venkatramanan, A. Warren, B. Lewis, J. Crow, R. Singh, A. Lorentz, D. Toney, M. Marathe, Wastewater-based epidemiology for covid-19 surveillance and beyond: A survey (2024). URL https://arxiv.org/abs/2403.15291

[8] P. M. D’Aoust, T. E. Graber, E. Mercier, D. Montpetit, I. Alexandrov, N. Neault, A. T. Baig, J. Mayne, X. Zhang, T. Alain, M. R. Servos, N. Srikanthan, M. MacKenzie, D. Figeys, D. Manuel, P. Jüni, A. E. MacKenzie, R. Delatolla, Catching a resurgence: Increase in sars-cov-2 viral rna identified in wastewater 48 h before covid-19 clinical tests and 96 h before hospitalizations, Science of The Total Environment 770 (2021) 145319. 10.1016/j.scitotenv.2021.145319. URL https://www.sciencedirect.com/science/article/pii/S0048969721003867

[9] V. Pileggi, J. Shurgold, J. S. Sun, M. I. Yang, E. A. Edwards, H. Peng, A. Tehrani, K. A. Gilbride, C. J. Oswald, R. Wijayasri, D. Al-Bargash, R. Stuart, Z. Khansari, M. Raby, J. Thomas, T. Fletcher, A. Simhon, Quantitative trend analysis of sars-cov-2 rna in municipal wastewater exemplified with sewershed-specific covid-19 clinical case counts, ACS ES&T Water 2 (11) (2022) 2070–2083. doi:10.1021/acsestwater.2c00058.

[10] P. M. D’Aoust, N. Hegazy, N. T. Ramsay, M. I. Yang, H. A. Dhiyebi, E. Edwards, M. R. Servos, G. Ybazeta, M. Habash, L. Goodridge, A. Poon, E. Arts, R. S. Brown, S. J. Payne, A. Kirkwood, D. Simmons, J.-P. Desaulniers, B. Ormeci, C. Kyle, D. Bulir, T. Charles, R. M. McKay, K. Gilbride, C. Oswald, H. Peng, V. Pileggi,M. L. Wang, A. Tong, D. Orellano, A. Adebiyi, M. Advani, S. Agboola, D. Andino, H. Aqeel, Y. Badlani, L. C. Bitter, L. Bragg, J. Brasset-Gorny, P. Breadner, S. Brown, R. Chan, B. Channa, J. Chen, R. Corchis-Scott, M. Cranney, H. Dang, N. Danna, R. Dawe, C. DeGroot, T. de Melo, H. Dhiyebi, J. Donovan, W. Eid, I. Ellmen, J. A. Farah, F. Farahbakhsh, M. Fuzzen, T. Garant, Q. Geng, A. Gedge, A. Gere, R. Gibson, K. Gilbride, E. Goitom, Q. C. Gong, T. Graber, A. Hamilton, B. Haskell, S. Hayat, H. Ho, Y. Hungwe, H. Ikert, G. Islam, D. Joseph, I. Khan, R. Kibbee, J. Knapp, J. Knockleby, S.-H. Kwon, O. U. Lawal, L. Lomheim, R. M. McKay, R. Menon, E. Mercier, Z. Miller, A. M. Mloszewska, A. Mohammadiankia, S. Naik, D. Nash, A. Ng, A. Olabode, B. Ormeci, A. Overton, G. J. Pabon, V. Paramananthasivam, J. Pardy, V. R. Parreira, L. Pisharody, S. Prasla, M. Precious, F. Rizvi, M. Santilli, H. Sarvi, M. Servos, D. Siemon, C. Sing-Judge, N. Srikanthan, S. Stephenson, J. S. Sun, E. Susilawati, A. Tehrani, O. Thakali, S. Wan, M. Wellman, K. Williams, I. Yang, E. Zeeb, E. M. Renouf, C. T. DeGroot, R. Delatolla, W. Consortium, Sars-cov-2 viral titer measurements in ontario, canada wastewaters throughout the covid-19 pandemic, Scientific Data 11 (1) (2024) 656. doi:10.1038/s41597-024-03414-w. URL 10.1038/s41597-024-03414-w

[11] Ontario Agency for Health Protection and Promotion (Public Health Ontario), Technical notes: Ontario respiratory virus tool (June 2024).

[12] Public Health Agency of Canada, Wastewater monitoring dashboard, Accessed January, 30, 2025 (June 2024). URL https://health-infobase.canada.ca/wastewater/about.html

[13] US Centers for Disease Control and Prevention, About Wastewater Data, NWSS, Accessed June 23, 2024 (May 2024).URL https://www.cdc.gov/nwss/about-data.html

[14] National Institute for Public Health and the Environment, Weekly Coronovirus SARS-CoV-2 Figures (December 2024). URL https://www.rivm.nl/en/coronavirus-covid-19/current/weekly-update

[15] Federal Envirorment Agency and Robert Koch Institut and Amelag and Umbwelt Bundesamt, Technical Guide Part 4 for Wastewater Surveillance – Data Processing, Accessed December 6, 2024 (2024). URL https://www.rki.de/EN/Content/Institute/DepartmentsUnits/InfDiseaseEpidem/Div32/WastewaterSurveillance/Guideline-4.pdf?blob=publicationFile

[16] Ministry for the Ecological Transition and Ministry of Health, Microbiological Control in Wastewater as an Early Epidemiological Indicator of COVID-19 Spread. Biweekly Report., Accessed December 11, 2024 (2024). URL https://www.miteco.gob.es/content/dam/miteco/es/agua/temas/concesiones-y-autorizaciones/vertidos-de-aguas-residuales/alerta-temprana-covid19/informes-actualizados/pdfs-informes-semanales/Resultados-Q29.pdf

[17] M. D. Damien, C. Galey, H. De Valk, F. Golliot, Virological surveillance of SARS-CoV-2 in wastewater in France: protocol for implementation from a public health perspective, Epidemiological section. Saint-Maurice: Santé publique France (March 2022).

[18] G. La Rosa, P. Mancini, G. Bonanno Ferraro, C. Veneri, M. Iaconelli, L. Bonadonna, L. Lucentini, E. Suffredini, Sars-cov-2 has been circulating in northern italy since december 2019: Evidence from environmental monitoring, Science of The Total Environment 750 (2021) 141711. 10.1016/j.scitotenv.2020. 141711. URL https://www.sciencedirect.com/science/article/pii/S0048969720352402

[19] A.-M. Hokajärvi, A. Rytkönen, A. Tiwari, A. Kauppinen, S. Oikarinen, K.-M. Lehto, A. Kankaanpää, T. Gunnar, H. Al-Hello, S. Blomqvist, I. T. Miettinen, C. Savolainen-Kopra, T. Pitkänen, The detection and stability of the sars-cov-2 rna biomarkers in wastewater influent in helsinki, finland, Science of The Total Environment 770 (2021) 145274. 10.1016/j.scitotenv.2021.145274. URL https://www.sciencedirect.com/science/article/pii/S0048969721003405

[20] J. S. Huisman, J. Scire, L. Caduff, X. Fernandez-Cassi, P. Ganesanandamoorthy, A. Kull, A. Scheidegger, E. Stachler, A. B. Boehm, B. Hughes, A. Knudson, A. Topol, K. Wigginton, M. K. Wolfe, T. Kohn, C. Ort, T. Stadler, T. R. Julian, Wastewater-Based Estimation of the Effective Reproductive Number of SARS-CoV-2, Environ Health Perspect 130 (May 2022). URL 10.1289/EHP10050

[21] Belgian Institute for Health (SCiensano), CoVWWSurv - National wastewater-based epidemiological surveillance, Accessed February 3, 2025 (2025). URL https://www.sciensano.be/en/projects/national-wastewater-based-epidemiological-surveillance

[22] The Portuguese Environment Agency (APA), Wastewater as the means of improving the response to new Covid outbreaks, Accessed February 3, 2025 (2025). URL https://www.adp.pt/en/news/?id=69&idn=417

[23] Statens Serum Institute, National surveillance of SARS-CoV-2 in wastewater, Accessed February 3, 2025 (2025). URL https://en.ssi.dk/surveillance-and-preparedness/surveillance-in-denmark/covid-19/national-surveillance-of-sars-cov-2-in-wastewater

[24] Australian Government Department of Health and Aged Care, SARS-CoV-2 Wastewater Surveillance CDNA National Strategy, accessed February 3, 2025 (2023). URL https://www.health.gov.au/resources/publications/sars-cov-2-wastewater-surveillance-cdna-national-strategy

[25] Ontario MECP, Ontario Clean Water Agency, MECP Wastewater Surveillance Initiative, Protocol for Evaluations of RT-qPCR Performance Characteristics (ISBN 978–1-4868-5481-3 Internal Laboratory Protocol), iSBN 978-1- 4868-5481-3 Internal Laboratory Protocol (August 2021).

[26] M. Fuzzen, N. B. Harper, H. A. Dhiyebi, N. Srikanthan, S. Hayat, L. M. Bragg, S. W. Peterson, I. Yang, J. Sun, E. A. Edwards, J. P. Giesy, C. S. Mangat, T. E. Graber, R. Delatolla, M. R. Servos, An improved method for determining frequency of multiple variants of sars-cov-2 in wastewater using qpcr assays, Science of The Total Environment 881 (2023) 163292. 10.1016/j.scitotenv.2023.163292. URL https://www.sciencedirect.com/science/article/pii/S0048969723019113

[27] PHES-ODM or ODM, The public health environmental surveillance open data model, version 2.2.3 (February 2024). URL https://github.com/Big-Life-Lab/PHES-ODM

[28] R Core Team, R: A Language and Environment for Statistical Computing, R Foundation for Statistical Computing, Vienna, Austria (2023). URL https://www.R-project.org/

[29] Public Health Ontario, Ontario COVID-19 Hospital Admissions and Deaths by Age from Waves 1-7, King’s Printer for Ontario (January 2023). URL https://www.publichealthontario.ca/-/media/Documents/nCoV/epi/2023/01/covid-19-hospital-admissions-deaths-age-epi-summary.pdf?rev=a65fe9c2cda34474af3011e919d057f8&sc_lang=en

[30] Land Information Ontario, Ministry of Health Public Health Unit Boundary, MOH PHU Boundary(January 2024). URL https://www.arcgis.com/home/item.html?id=c2fa5249b0c2404ea8132c051d934224

[31] RStudio Team, RStudio: Integrated Development Environment for R, RStudio, PBC, Boston, MA (2021). URL http://www.rstudio.com/

[32] D. H. Ogle, J. C. Doll, A. P. Wheeler, A. Dinno, FSA: Simple Fisheries Stock Assessment Methods, r package version 0.9.6 (2025). URL https://CRAN.R-project.org/package=FSA

[33] J. B. Arnold, ggthemes: Extra Themes, Scales and Geoms for ‘ggplot2’, r package version 5.0.0 (2023). URL https://CRAN.R-project.org/package=ggthemes

[34] B. Auguie, gridExtra: Miscellaneous Functions for “Grid” Graphics, r package version 2.3 (2017). URL https://CRAN.R-project.org/package=gridExtra

[35] G. Grolemund, H. Wickham, Dates and times made easy with lubridate, Journal of Statistical Software 40 (3) (2011) 1–25. URL https://www.jstatsoft.org/v40/i03/

[36] F. E. Harrell Jr, Hmisc: Harrell Miscellaneous, r package version 5.1-1 (2023). URL https://CRAN.R-project.org/package=Hmisc

[37] F. E. Harrell Jr, rms: Regression Modeling Strategies, r package version 6.7-1 (2023). URL https://CRAN.R-project.org/package=rms

[38] S. Højsgaard, U. Halekoh, doBy: Groupwise Statistics, LSmeans, Linear Estimates, Utilities, r package version 4.6.20 (2023). URL https://CRAN.R-project.org/package=doBy

[39] A. Jelenko, P. Punco, neatRanges: Tidy Up Date/Time Ranges, r package version 0.1.4 (2022). URL https://CRAN.R-project.org/package=neatRanges

[40] A. Kassambara, ggpubr: ‘ggplot2’ Based Publication Ready Plots, r package version 0.6.0 (2023). URL https://CRAN.R-project.org/package=ggpubr

[41] J. Landis, ggside: Side Grammar Graphics, r package version 0.2.3 (2023). URL https://CRAN.R-project.org/package=ggside

[42] F. Lindgren, fmesher: Triangle Meshes and Related Geometry Tools, r package version 0.1.5 (2023). URL https://CRAN.R-project.org/package=fmesher

[43] D. Makowski, D. Lüdecke, I. Patil, R. Thériault, M. S. Ben-Shachar, B. M. Wiernik, Automated results reporting as a practical tool to improve reproducibility and methodological best practices adoption, CRAN (2023). URL https://easystats.github.io/report/

[44] K. Müller, H. Wickham, tibble: Simple Data Frames, r package version 3.2.1 (2023). URL https://CRAN.R-project.org/package=tibble

[45] D. Murdoch, D. Adler, rgl: 3D Visualization Using OpenGL, r package version 1.2.8 (2023). URL https://CRAN.R-project.org/package=rgl

[46] P. Murrell, RGraphics: Data and Functions from the Book R Graphics, Third Edition, r package version 3.0–2 (2020). URL https://CRAN.R-project.org/package=RGraphics

[47] E. Neuwirth, RColorBrewer: ColorBrewer Palettes, r package version 1.1-3 (2022). URL https://CRAN.R-project.org/package=RColorBrewer

[48] J. Ooms, Kornel Lesiński, Authors of the dependency Rust crates, gifski: Highest Quality GIF Encoder, r package version 1.12.0-2 (2023). URL https://CRAN.R-project.org/package=gifski

[49] E. Pebesma, R. Bivand, Spatial Data Science: With applications in R, Chapman and Hall/CRC, 2023. doi: 10.1201/9780429459016. URL https://r-spatial.org/book/

[50] E. Pebesma, Simple Features for R: Standardized Support for Spatial Vector Data, The R Journal 10 (1) (2018) 439–446. doi:10.32614/RJ-2018-009. URL 10.32614/RJ-2018-009

[51] B. Schloerke, D. Cook, J. Larmarange, F. Briatte, M. Marbach, E. Thoen, A. Elberg, J. Crowley, GGally: Extension to ‘ggplot2’, r package version 2.2.1 (2024). URL https://CRAN.R-project.org/package=GGally

[52] M. Tennekes, tmaptools: Thematic Map Tools, r package version 3.1-1 (2021). URL https://github.com/mtennekes/tmaptools

[53] G. R. Warnes, B. Bolker, T. Lumley, A. Magnusson, B. Venables, G. Ryodan, S. Moeller, gtools: Various R Programming Tools, r package version 3.9.5 (2023). URL https://CRAN.R-project.org/package=gtools

[54] H. Wickham, Reshaping data with the reshape package, Journal of Statistical Software 21 (12) (2007). URL https://www.jstatsoft.org/v21/i12/

[55] H. Wickham, The split-apply-combine strategy for data analysis, Journal of Statistical Software 40 (1) (2011) 1–29. URL https://www.jstatsoft.org/v40/i01/

[56] H. Wickham, ggplot2: Elegant Graphics for Data Analysis, Springer-Verlag New York, 2016. URL https://ggplot2.tidyverse.org

[57] H. Wickham, forcats: Tools for Working with Categorical Variables (Factors), r package version 1.0.0 (2023). URL https://CRAN.R-project.org/package=forcats

[58] H. Wickham, stringr: Simple, Consistent Wrappers for Common String Operations, r package version 1.5.1 (2023). URL https://CRAN.R-project.org/package=stringr

[59] H. Wickham, M. Averick, J. Bryan, W. Chang, L. D. McGowan, R. François, G. Grolemund, A. Hayes, L. Henry, J. Hester, M. Kuhn, T. L. Pedersen, E. Miller, S. M. Bache, K. Müller, J. Ooms, D. Robinson, D. P. Seidel, V. Spinu, K. Takahashi, D. Vaughan, C. Wilke, K. Woo, H. Yutani, Welcome to the tidyverse, Journal of Open Source Software 4 (43) (2019) 1686. doi:10.21105/joss.01686.

[60] H. Wickham, J. Bryan, readxl: Read Excel Files, r package version 1.4.3 (2023). URL https://CRAN.R-project.org/package=readxl

[61] H. Wickham, J. Bryan, M. Barrett, A. Teucher, usethis: Automate Package and Project Setup, r package version 2.2.2 (2023). URL https://CRAN.R-project.org/package=usethis

[62] H. Wickham, R. François, L. Henry, K. Müller, D. Vaughan, dplyr: A Grammar of Data Manipulation, r package version 1.1.4 (2023). URL https://CRAN.R-project.org/package=dplyr

[63] H. Wickham, L. Henry, purrr: Functional Programming Tools, r package version 1.0.2 (2023). URL https://CRAN.R-project.org/package=purrr

[64] H. Wickham, J. Hester, J. Bryan, readr: Read Rectangular Text Data, r package version 2.1.5 (2024). URL https://CRAN.R-project.org/package=readr

[65] H. Wickham, J. Hester, W. Chang, J. Bryan, devtools: Tools to Make Developing R Packages Easier, r package version 2.4.5 (2022). URL https://CRAN.R-project.org/package=devtools

[66] H. Wickham, T. L. Pedersen, D. Seidel, scales: Scale Functions for Visualization, r package version 1.3.0 (2023). URL https://CRAN.R-project.org/package=scales

[67] H. Wickham, D. Vaughan, M. Girlich, tidyr: Tidy Messy Data, r package version 1.3.1 (2024). URL https://CRAN.R-project.org/package=tidyr

[68] C. O. Wilke, cowplot: Streamlined Plot Theme and Plot Annotations for ‘ggplot2’, r package version 1.1.1 (2020). URL https://CRAN.R-project.org/package=cowplot

[69] Y. Xie, bookdown: Authoring Books and Technical Documents with R Markdown, Chapman and Hall/CRC, Boca Raton, Florida, 2016. URL https://bookdown.org/yihui/bookdown

[70] Y. Xie, Implementing Reproducible Computational Research knitr: A Comprehensive Tool for Reproducible Research in R, Chapman and Hall/CRC, 2014, iSBN 978-1466561595.

[71] A. Zeileis, G. Grothendieck, zoo: S3 infrastructure for regular and irregular time series, Journal of Statistical Software 14 (6) (2005) 1–27. doi:10.18637/jss.v014.i06.

[72] A. Rabe, S. Ravuri, E. Burnor, J. A. Steele, R. S. Kantor, S. Choi, S. Forman, R. Batjiaka, S. Jain, T. M. León, D. J. Vugia, A. T. Yu, Correlation between wastewater and covid-19 case incidence rates in major california sewersheds across three variant periods, Journal of Water and Health (2023). URL 10.2166/wh.2023.173

[73] Ontario Agency for Health Protection and Promotion (Public Health Ontario), Epidemiologic summary: Sars-cov-2 whole genome sequencing in ontario: Queen’s printer for ontario (December 2022). URL https://www.publichealthontario.ca/-/media/Documents/nCoV/Archives/Genome/2022/12/SARS-CoV-2-genomic-surveillance-report-2022-12-28.pdf?rev=bbda499b034540bcb32f0b0215891031&sc_lang=en

[74] L. Cheng, H. A. Dhiyebi, M. Varia, K. Atanas, N. Srikanthan, S. Hayat, H. Ikert, M. Fuzzen, C. Sing-Judge, Y. Badlani, E. Zeeb, L. M. Bragg, R. Delatolla, J. P. Giesy, E. Gilliland, M. R. Servos, Omicron covid-19 case estimates based on previous sars-cov-2 wastewater load, regional municipality of peel, ontario, canada, Emerging Infectious Diseases (2023). URL 10.3201/eid2908.221580

[75] Ministry of Health, Updated Eligibility for PCR Testing and Case and Contact Management Guidance in Ontario (December 2021). URL https://news.ontario.ca/en/backgrounder/1001387/updated-eligibility-for-pcr-testing-and-case-and-contact-management-guidance-in-ontario

[76] Ministry of Health, Ontario expands covid-19 testing to pharmacies (September 2020). URL https://news.ontario.ca/en/release/58492/ontario-expands-covid-19-testing-to-pharmacies

[77] Ministry of Health, Ontario updates covid-19 testing guidelines (September 2020). URL https://news.ontario.ca/en/statement/58507/ontario-updates-covid-19-testing-guidelines

[78] Office of the Premier, Ontario Implementing Additional Public Health and Testing Measures to Keep People Safe (October 2020). URL https://news.ontario.ca/en/release/58645/ontario-implementing-additional-public-health-and-testing-measures-to-keep-people-safe

[79] Ottawa Public Health, Seasonal Respiratory Infections and Enteric Outbreaks Surveillance Dashboard (February 2025). URL https://www.ottawapublichealth.ca/en/reports-research-and-statistics/flu-report.aspx

[80] Y. Wang, Q. Di, Modifiable areal unit problem and environmental factors of covid-19 outbreak, Science of The Total Environment 740 (2020) 139984. 10.1016/j.scitotenv.2020.139984. URL https://www.sciencedirect.com/science/article/pii/S004896972033504X

[81] Élisabeth Mercier, P. M. D’Aoust, W. Eid, N. Hegazy, P. Kabir, S. Wan, L. Pisharody, E. Renouf, S. Stephenson, T. E. Graber, A. E. MacKenzie, R. Delatolla, Sewer transport conditions and their role in the decay of endogenous sars-cov-2 and pepper mild mottle virus from source to collection, International Journal of Hygiene and Environmental Health 263 (2025) 114477. 10.1016/j.ijheh.2024.114477. URL https://www.sciencedirect.com/science/article/pii/S1438463924001585

[82] Y. Ye, R. M. Ellenberg, K. E. Graham, K. R. Wigginton, Survivability, partitioning, and recovery of enveloped viruses in untreated municipal wastewater, Environmental Science & Technology 50 (10) (2016) 5077–5085. doi:10.1021/acs.est.6b00876. URL 10.1021/acs.est.6b00876

